# Transdiagnostic Clinical Features Delineate Trajectories of Serious Mental Illness

**DOI:** 10.1101/2022.08.20.22279007

**Authors:** Juan F. De la Hoz, Alejandro Arias, Susan K. Service, Mauricio Castaño, Ana M. Diaz-Zuluaga, Janet Song, Cristian Gallego, Sergio Ruiz-Sánchez, Javier I Escobar, Alex A. T. Bui, Carrie E. Bearden, Victor Reus, Carlos Lopez-Jaramillo, Nelson B. Freimer, Loes M. Olde Loohuis

**Affiliations:** Center for Neurobehavioral Genetics, Semel Institute for Neuroscience and Human Behavior, David Geffen School of Medicine, University of California Los Angeles, Los Angeles, USA; Department of Mental Health and Human Behavior, University of Caldas, Manizales, Colombia; Department of Psychiatry, University of Antioquia, Medellín, Colombia; Global Health, Robert Stempel School of Public Health and Social Work, Florida International University, Miami, USA; Department of Radiological Sciences, University of California Los Angeles, California, USA; Department of Psychiatry and Biobehavioral Sciences, University of California San Francisco, San Francisco, USA

## Abstract

**Background:** Electronic health record (EHR) databases, increasingly available in low- and middle-income countries (LMIC), provide an opportunity to study transdiagnostic features of serious mental illness (SMI) and delineate illness trajectories using clinical data.

**Aims:** Characterize transdiagnostic features and diagnostic trajectories of SMI using structured and unstructured data from an EHR database in an LMIC institution.

**Methods:** We conducted a retrospective cohort study using EHR data from 2005-2022 at Clínica San Juan de Dios Manizales, a specialized mental health facility in Caldas, Colombia. We included 22,447 patients treated for schizophrenia (SCZ), bipolar disorder (BD), severe or recurrent major depressive disorder (MDD). We extracted diagnostic codes, clinical notes, and healthcare use data from the EHR database. Using natural language processing, we analyzed the frequency of suicidality and psychosis across SMI diagnoses. Using the diagnostic trajectories, we studied patterns of diagnostic switching and accumulation of comorbidities. Mixed-effect logistic regression was used to assess factors influencing diagnostic stability.

**Results:** High frequencies of suicidality and psychosis were observed across diagnoses of SCZ, BD, and MDD. Most SMI patients (64%) received multiple diagnoses over time, including switches between primary SMI diagnoses (19%), diagnostic comorbidities (30%), or both (15%). Predictors of diagnostic switching included mentions of delusions in clinical notes (OR=1.50, p=2e-18), prior diagnostic switching (OR=4.02, p=3e-250), and time in treatment, independent of age (log of visit number; OR=0.56, p=5e-66). Over 80% of patients reached diagnostic stability within six years of their first record.

**Conclusions:** This study demonstrates that integrating structured and unstructured EHR data can reveal clinically relevant, transdiagnostic patterns in SMI, including early predictors of disease trajectories. Our findings underscore the potential of EHR-based tools to aid etiological research and the development of personalized treatment strategies, particularly in LMIC.

## Introduction

Examination of disease trajectories through longitudinal clinical observation of symptoms led to the development of modern classification systems for mental disorders, which differentiate categories of serious mental illness (SMI), including schizophrenia (SCZ), bipolar disorder (BD), and severe major depressive disorder (MDD). While such classification systems advocate a parsimonious approach in which patients are assigned unique diagnoses, this conflicts with the clinical reality that many features of psychiatric illness (such as suicidality or psychosis) are present across several diagnoses ^1^. Furthermore, while traditional classification systems reflect a longitudinal perspective, current research on SMI relies primarily on cross-sectional assessments, in which the only available trajectory information is supplied by patient recall ^2–4^. This lack of detailed longitudinal data may be a factor contributing to the heterogeneity observed in studies based on current SMI categories ^5^, notably in cross-disorder genetic analyses ^4^. Additionally, while cross-sectional data may support the notion that each patient may be characterized according to a unique diagnosis, it may take several years after initial presentations for most patients with SMI to achieve a stable diagnosis ^6,7^.

Recent studies using longitudinal data collected from participants in national registries ^8,9^, precision health initiatives ^10^, and birth cohorts ^11^ have begun to identify transdiagnostic risk factors and to describe patterns of variation across diagnoses over time ^8–11^. These resources, which are mainly limited to upper-income countries (UIC), typically contain only sparse data for individual clinical features, such as symptoms and behaviors. In contrast, electronic health record (EHR) data, available in both UIC and in many low- and middle–income countries (LMIC), may contain extensive descriptions of such clinical features during the periods when patients experience them. As we demonstrate here for an institution located in an LMIC, EHR databases thus facilitate investigations of features that are important both transdiagnostically and longitudinally, and that may predict clinically important outcomes, such as the onset of psychosis ^12^, the occurrence of suicidal behaviors ^13^, or the stability of clinical diagnoses ^14,15^.

## Methods

### EHR database

The Clínica San Juan de Dios in Manizales, Colombia (CSJDM), provides comprehensive mental healthcare to the one million inhabitants of the department (state) of Caldas ^16^. For this study, we extracted structured EHR data collected between 2005 and 2022, including demographic information; duration, type, and site of visits (inpatient, outpatient, or emergency department); diagnostic codes (ICD-10); and unstructured data, consisting of free-text from clinical notes. For our analyses, we included all patients with at least one clinical note in their EHR, and excluded patients with missing gender information. We excluded visits that were outside the age range of 4-90, without a valid diagnostic code, or with primary diagnostic codes outside of chapter V of the ICD-10 (Mental, Behavioral, and Neurodevelopmental Disorders categories), Supplementary Figure 1.

### ICD-10 codes extraction and cohort definition

Following each visit to the hospital, a patient is assigned a single primary ICD-10 diagnosis by their treating psychiatrist, generating a time-stamped sequence of diagnoses. We extracted this sequence for every patient and selected for analyses patients who had at least one primary diagnosis of SMI, defined here as BD (F301, F302, F310, F311, F312, F313, F314, F315, F316, F317), severe/recurrent MDD (F322, F323, F331, F332, F333, F334), SCZ (F20X), and other chronic psychoses (delusional disorder; F22X. schizoaffective disorder; F25X) (Supplementary Table 1). In total, this cohort includes 22,447 patients with 157,003 visits (Supplementary Figure 1B).

### Reliability of the current ICD-10 diagnosis and its association with clinical features

We quantified the reliability of the current ICD-10 diagnosis by comparing them to those obtained through a complete manual chart review (Supplementary Note 1). We then used a Spanish-language natural language processing (NLP) algorithm to extract clinical features from free-text notes (Supplementary Note 2). Specifically, we focused on four transdiagnostic features that are routinely assessed in clinical practice: suicide attempts, suicidal ideation, delusions, and hallucinations. We tested the relationship between these four clinical features, current diagnosis, and gender in SMI patients with at least two recorded clinical notes (Supplementary Figure 1C). Association tests for each feature were performed using logistic regression, adjusting for the length of patients’ records and history of hospitalization. We used this logistic modeling framework to evaluate the effect of co-occurring clinical features by adding, for each feature, the presence of the three remaining features. We also tested for interactions between gender and the second co-occurring feature by including an interaction term (Supplementary Note 3).

### Diagnostic trajectories

To describe the diversity of diagnostic trajectories observed in the EHR database, we used the sequence of primary ICD-10 diagnoses extracted above. We defined two types of diagnostic changes: diagnostic switches and the addition of comorbidities. We use the term *diagnostic switches* to refer to changes between two psychiatric diagnoses that cannot, by definition, be held at the same time; specifically, the diagnoses in the ICD-10 F2 and F3 chapters (psychotic and mood disorders, respectively; Supplementary Note 4 and Supplementary Table 2). By contrast, we use the term diagnostic *comorbidities* to refer to all other combinations of ICD-10 codes; comorbid psychiatric diagnoses can accumulate over time, without limit. We used this definition of diagnostic trajectories in patients with at least three recorded visits (Supplementary Figure 1D) to estimate the proportion of patients with diagnostic switches, recorded comorbidities, or both.

### Factors affecting diagnostic stability

We explored factors contributing to visit-to-visit diagnostic stability. First, we used a mixed-effect logistic regression to estimate the probability of switching diagnoses as a function of time, accounting for repeated patient observations. We measured time as the number of visits, and separately, as the number of years since the first visit. Then, we expanded this model to include ten additional factors: (i) patient’s gender and (ii) age, (iii) current primary diagnosis, (iv) inpatient status, (v-viii) the four NLP-derived clinical features, (ix) receiving a “not otherwise specified” (NOS) code, and (x) previous diagnostic switching (details in Supplementary Note 5). An NOS code indicates diagnostic uncertainty in cases of atypical or confusing patient presentations or when temporal criteria are not yet met ^17^. As we expect NOS codes to be associated with a higher degree of diagnostic instability than other codes, we consider them to serve as a positive control.

To evaluate the possibility that clinical features extracted from the notes at a given visit *anticipate* specific diagnostic changes recorded in future visits, we tested whether psychosis features (delusions and hallucinations) predict a subsequent application of ICD-10 codes specifying psychotic features in diagnoses of BD or MDD (Supplementary Note 6; Supplementary Figure 1E).

Finally, to estimate the time it takes to reach diagnostic stability, we analyzed patients with records of 10 years or longer (Supplementary Figure 1F). We defined stability as the absence of future diagnostic switches. For each year of follow-up, we calculated the percentage of patients reaching stability by that time. We also identified patients with high levels of diagnostic instability as those who had five or more diagnostic switches and at least one of them occurred after five years of illness.

### Significance thresholds

We applied Bonferroni correction for multiple testing in all our analyses. Model details with corresponding significance thresholds are described in Supplementary Notes 3, 5, and 6.

### Ethical approval

This study was approved by the Institutional Review Board (IRB) of UCLA, the Comité de Ética del Instituto de Investigaciones Médicas at Universidad de Antioquia (UdeA), and the Comité de Bioética at CSJDM.

## Results

### Study sample

As of June 2022, the CSJDM EHR included 157,003 visits from 22,447 patients who were assigned an SMI diagnosis at any point in their records (Supplementary Figure 1B). The demographic and clinical characteristics of this sample are described in Supplementary Table 8.

### Transdiagnostic characterization of features extracted from EHR notes

We found that compared to manual chart reviews, patients’ current ICD-10 diagnoses of MDD, BD, and SCZ were highly accurate (accuracy of 0.90, 0.88, and 0.95, respectively, Supplementary Table 3). Each of the four NLP-extracted features (suicidal ideation, suicide attempt, delusions, and hallucinations) occurred in all of the SMI diagnoses, stratified by gender, at frequencies above 5%, demonstrating their transdiagnostic quality (Figure 1A).

**Figure 1.**
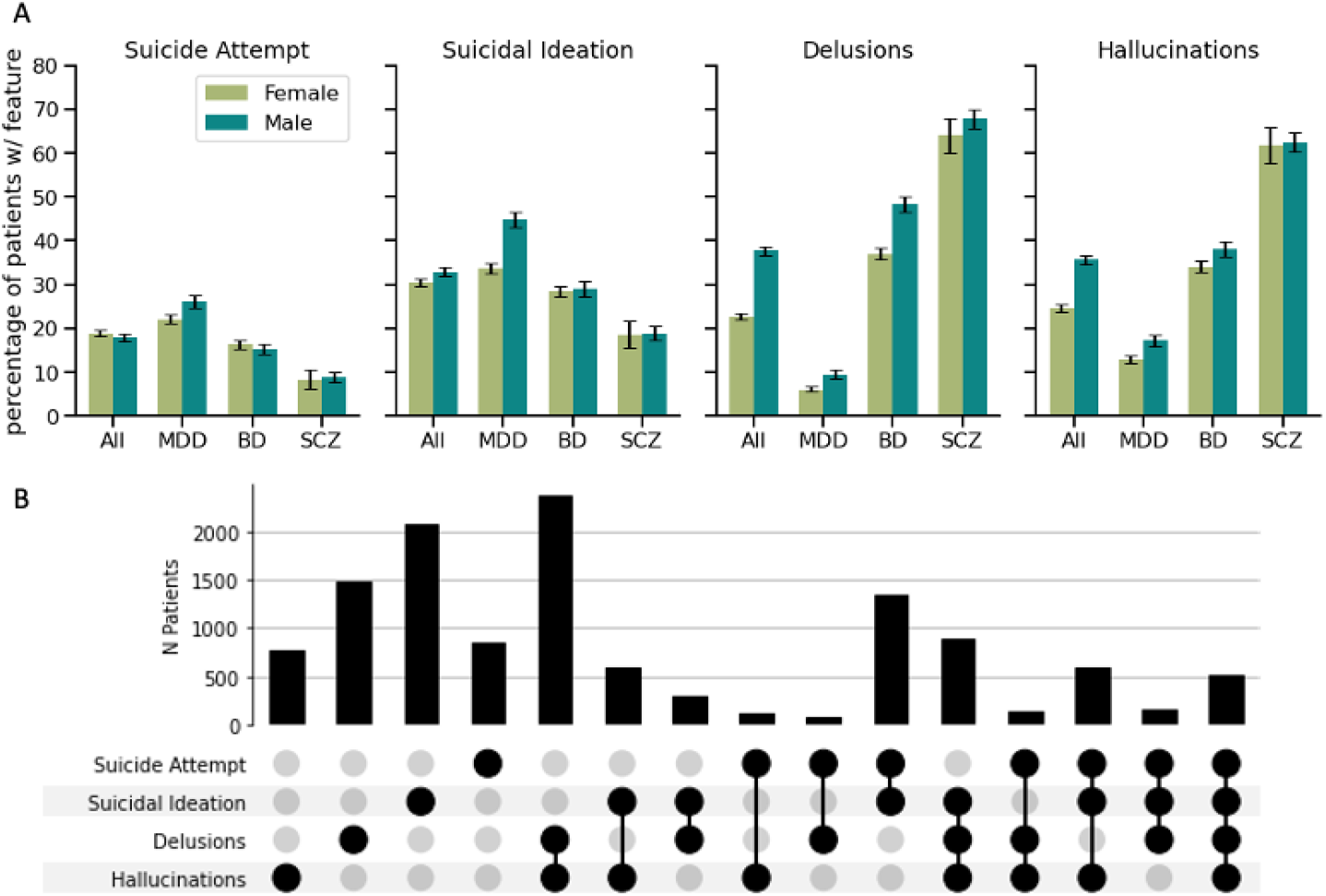
Transdiagnostic characterization and co-occurrence of clinical features extracted from EHR notes. A) Proportion of patients with each of the four features stratified by primary diagnosis. B) Number of patients with co-occurrence of 2, 3, or 4 clinical fetures. All data in these plots are limited to patients with at least two EHR notes.

Suicidal ideation is, in most studies, more frequent in females than males ^18–20^. We observed the reverse pattern in our dataset (OR=0.84, p=8.42e^-7^, Supplementary Tables 9 and 10, after correcting for diagnoses, inpatient history, and number of visits), a finding largely driven by the relatively low frequency of suicidal ideation in females compared to males with MDD (34% vs. 45%, interaction OR=0.65, p=4.2e^-9^). Rates of suicide attempt, in our dataset, were similar in both genders. Across the SMI diagnoses, as expected ^21^, the two types of psychotic features also showed a reduced frequency in females compared to males (across all diagnoses, OR=0.67, p=1.99e^-22^ delusions; and OR=0.88, p=5.9e^-4^ hallucinations). This pattern was further evident in each of the diagnoses considered separately; in contrast, the preponderance of previous studies have found no significant difference between males and females in the frequency of psychotic symptoms in MDD or BD ^21^.

The co-occurrence of the four features is displayed in Figure 1B. As expected, the two suicide-related features tended to co-occur as did the two psychotic features. The mention of hallucinations increased the likelihood of mention of suicidal features, and vice versa (OR between 1.29-2.05, p < 2.89e^-7^), accounting for gender, diagnosis, inpatient history and number of visits (Supplementary Table 10). However, unexpectedly, the mention of delusions in the notes decreased the likelihood of notes mentioning either suicidal feature in the same patient and vice versa (OR between 0.59-0.62, p < 1.78e^-17^).

### Diverse diagnostic trajectories in SMI

We described diagnostic trajectories among SMI patients with at least three recorded visits (n=12,962; Supplementary Figure 1D). The majority (64%, Figure 2A) had multiple diagnoses recorded in their EHR, broken down as follows: 30% displayed comorbidities (orange bars; Supplementary Table 11), 19% displayed diagnostic switches (teal bars; Supplementary Figure 3), and 15% displayed both switches and comorbidities (purple bars). Early switches often involved a change from brief psychotic disorder or single-episode MDD to BD ^22^, while later switches frequently involved SCZ, BD and schizoaffective disorder ^23^ (Supplementary Figure 4).

**Figure 2:**
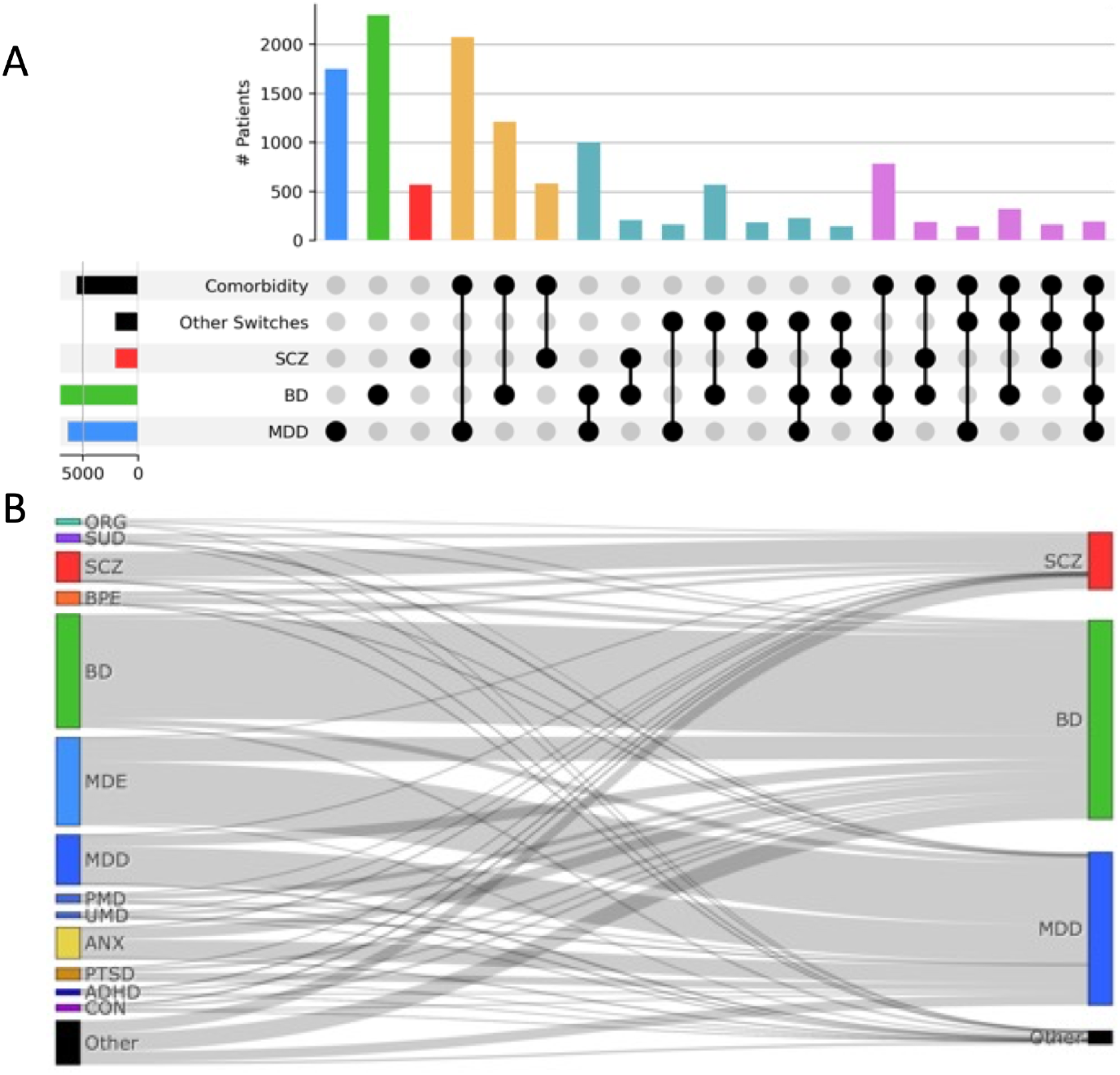
Disease trajectories of SMI in patients with at least three visits. A) UpSet plot presenting diagnostic switches (between SMI categories) and comorbidities (SMI and non-SMI categories). Patients with a single SMI diagnosis (blue, green, red, total n=4,620); a single SMI diagnosis and other comorbidities (orange n=3,955); multiple SMI diagnoses and no other comorbidities (teal n=2,468); multiple SMI diagnoses and other comorbidities (purple, n=1,919). Bars with n<100 are not shown. B) Sankey diagram of diagnostic trajectories. The left nodes represent the diagnosis given at the initial visit, and the right nodes represent the most recent SMI code. (Diagnostic switches within SMI are shown in Supplementary Figure SF3). ORG: Other mental disorders due to brain damage and dysfunction and to physical disease (F06), SUD: Mental and behavioral disorders due to multiple drug use and use of other psychoactive substances (F19), BPE: Acute and transient psychotic disorders (F23), MDE: Major Depressive Episode (F32), PMD: Persistent mood disorders (F34), UMD: Unspecified mood disorder (F39), ANX: Other anxiety disorders (F41), PTSD: Reaction to severe stress, and adjustment disorders (F43), ADHD: Hyperkinetic disorders (F90), CON: Conduct disorders (F91)

Some trajectories are comprised of diagnoses that are frequently paired, e.g., the diagnostic switch from MDD to BD (observed in 22% of current BD patients) or the comorbidity between MDD and Other Anxiety Disorders (observed in 28% of current MDD patients). We found that the majority of cases (58%) follow rare trajectories (occurring in fewer than 1% of patients). Altogether, we counted 3,149 unique trajectories.

### Clinical features, time, and other factors affecting diagnostic stability

We identified multiple factors that influenced diagnostic stability. Diagnostic switching was most frequent during the early stages of treatment. While 11.3% of the patients changed diagnosis on their second visit, this percentage decreased over the patient’s course of illness (Figure 3A; log10(k) OR=0.56, p-value 5e^-66^) and stabilized at around 4% after the tenth visit. Additional predictors of future diagnostic instability included the following observations at the current visit: a diagnostic switch from the previous visit (OR=4.02, p-value 3e^-250^; Supplementary Figure 5), an inpatient visit (OR=1.7, p-value 5e^-35^), an NOS diagnosis (OR=1.61, p-value 2e^-47^), and the presence of the clinical features delusions or hallucinations (OR=1.50 and 1.17, p-values 2e^-18^ and 3e^-4^, respectively). Predictors of future diagnostic stability included diagnoses of SCZ or BD compared to MDD (ORs=0.31 and 0.32; p-values <3e^-70^), male gender (OR=0.71, p-value 2e^-16^), and age (OR per decade=0.96, p-value 8e^-4^). The same pattern was observed when modeling switching by time rather than visit number (Supplementary Figure 5).

**Figure 3:**
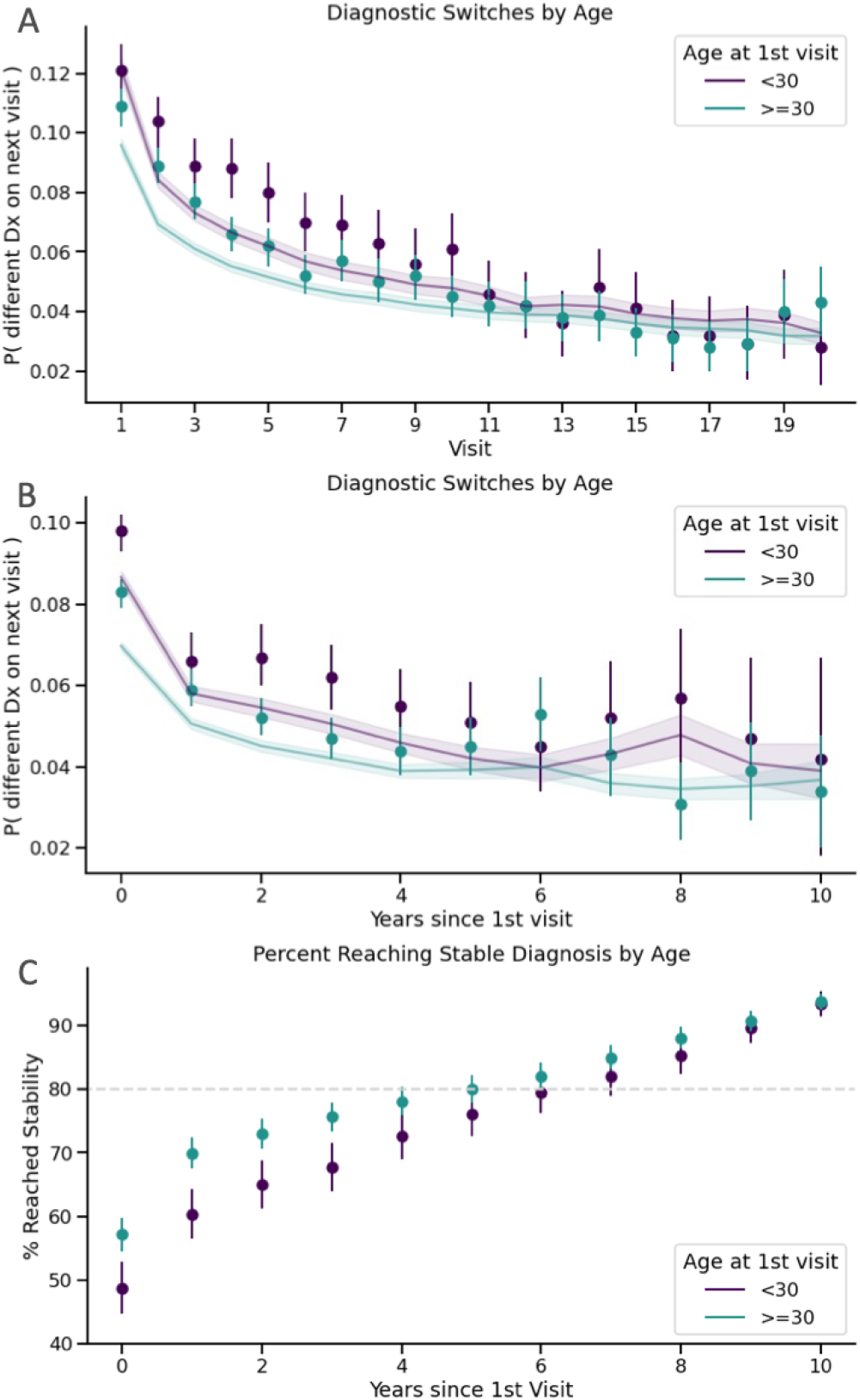
Diagnostic stability over time. A) At each visit k, the proportion of patients that will switch primary diagnosis code on their next visit k+1. Stratified by age groups: age at 1^st^ visit before and after 30 years. B) x-axis shows time since the first encounter instead of visit number. For every year, the observed proportion of visits that will have a diagnostic switch on the next visit. The solid line is the average probability of switching at any given visit during that year, as estimated by the model. Lines and shaded areas correspond to 95% confidence intervals. C) Proportion of patients by year, who have reached a stable diagnosis. N=1,952 patients with 10 or more years in the EHR. It takes 6 years for 80% of patients to reach a stable diagnosis.

### Reaching diagnostic stability

We identified 1,952 patients with over 10 years of EHR data. Of these, more than 80% reached a stable diagnosis within 6 years (Figure 3C). Additionally, 162 individuals showed high levels of diagnostic instability. In this group, switches between BD and SCZ were most common (Supplementary Table 12).

## Discussion

We leveraged EHR data spanning 17 years and encompassing over 20,000 patients from a large mental health facility in an LMIC to characterize transdiagnostic and longitudinal features of SMI. Previous studies describing the association of demographic variables and clinical features have varied greatly in methodology and scale ^18,19,21,24–27^. Our study stands out in four ways. First, we uniformly applied an NLP approach to clinical notes, to determine the presence or absence of four clinically important SMI features – suicidal ideation, suicide attempts, delusions, and hallucinations – detecting them at high frequencies across all SMI diagnoses. Second, we tested for gender differences across these features after adjusting for factors such as diagnosis, treatment duration, and age. Third, we characterized diagnostic trajectories in terms of diagnostic switches, the accumulation of comorbidities. And fourth, we evaluated the role of transdiagnostic features in predicting the stability of diagnoses.

### Transdiagnostic Features

The “gender paradox of suicide” – the observation of higher rates of suicidal ideation and suicide attempts in females compared to males, but higher rates of completed suicide in males – is well-documented globally, including in Latin America ^18,20,28–31^. Our observation of a higher rate of suicidal ideation in males compared to females was therefore unexpected and warrants further investigation. Regarding suicide attempts, we found approximately equal frequencies among males and females. This may reflect the high severity of patients in the EHR database; supporting this interpretation, a cross-national study reported a male excess for suicide attempts that were designated as “serious” ^32^. However, our current methodology has two important limitations. First, it does not distinguish between different levels of severity of suicide attempt, and second, it showed a lower recall for suicide attempts compared to other features, indicating that the prevalence reported here is likely an underestimate.

Psychotic symptoms have consistently been linked with higher suicidality rates across various SMI diagnoses ^25,33,34^. Interestingly, our study found opposite associations between suicidal features and specific psychotic symptoms – positive for hallucinations, negative for delusions. Our findings uniquely contribute to the existing literature, which typically does not differentiate between types of psychotic symptom. Future research will explore factors contributing to these observations, like substance use, and refine the extracted phenotypes for more detailed analyses, such as examining relationships between specific delusions and suicidal features.

Evidence has accumulated indicating a high degree of shared genetic risk across SMI diagnoses ^4^. It has been hypothesized that this shared risk may reflect genetic associations to transdiagnostic component phenotypes, but such phenotypes have rarely been assessed at a scale adequate to test this hypothesis. The results that we present here for suicidality and psychotic symptoms suggest that our approach for extracting transdiagnostic features from EHR notes may provide a general strategy for mounting well-powered genetic association studies of such phenotypes in cohorts ascertained for SMI, broadly. In future studies we plan to deploy such a strategy through investigations of additional EHR databases and inclusion of a more extensive set of clinical features.

### Diagnostic Trajectories

The continuous EHR record since 2005 at the CSJDM allowed us to analyze longitudinal trajectories of SMI. Consistent with previous research ^8,9,35,36^, we find that diagnostic instability is characteristic of the early stages of SMI, and that this contributes to a large diversity of disease trajectories. This diversity of trajectories underscores the complexity of SMI, and highlights the need to identify patterns relevant for understanding disease causation and informing clinical practice.

We found that most SMI patients in the CSJDM achieve diagnostic stability within 6 years, consistent with reports from UICs ^6,7^. Despite methodological differences, the overall alignment with previous registry studies ^8,9^ lends additional validation to our approach. However, our study’s unique contribution lies in the integration of features from clinical notes alongside diagnostic codes. This enabled us to delineate trajectories with greater granularity than typically available in registry data. We observed that clinical notes often documented psychotic features prior to the appearance of formal diagnostic codes for psychotic episodes, underscoring the potential for clinical notes to enhance predictive modeling in psychiatric care. Tools leveraging EHR data in predicting psychiatric outcomes are starting to emerge in UICs ^37,38^ – our work paves the way for future developments of this kind in LMICs.

Our results support the presumption that research classifications incorporating past and future trajectories at both symptom and diagnosis levels will lead to less heterogeneous categories than those that are based only on a ‘lifetime’ diagnosis ^39^. Prior studies have suggested that certain genetic risk profiles might contribute to specific SMI trajectories, such as polarity at the onset of BD ^40^ or conversion from non-psychotic to psychotic illness ^41–43^. Efforts to replicate and extend such findings, however, have been limited by variation in ascertainment strategies, reliance on patient recall ^40^, and small sample sizes ^41,43^. Centering genetic studies of SMI trajectories on EHR databases, such as that of the CSJDM, could provide a means to overcome these limitations; but as large-scale analyses of thousands of different disease trajectories appears impractical, it will be crucial to develop methods for reducing dimensionality by clustering patients with similar trajectories ^44^.

### Limitations

As described above, the limited granularity of clinical features extracted from free-text notes (e.g., we have not extracted specific types of delusions or the level of severity of suicide attempts) and the potential underestimation of true suicide attempts are limitations of this work. We are currently improving our NLP algorithms to address both limitations (e.g., by identifying instances of language that signifies intent to die), and simultaneously we are exploring approaches to expand our NLP toolset (e.g., by including a combination of pattern-based detection and Large Language Models ^45^). Along with this, a key limitation of using administrative data for research, including in this study, is the inability to differentiate between true diagnostic switches and variation in clinician subjectivity. Future studies involving extensive chart reviews at switch points could help evaluate this limitation.

## Conclusion

Our results demonstrate the utility of EHR databases for population-level research on SMI in an LMIC setting at unprecedented detail and scale. The availability of longitudinal EHRs enables the characterization of SMI trajectories over extended periods of time, while the use of NLP to uniformly phenotype patients across diagnoses enables investigation of transdiagnostic components of SMI. Extension of this approach could play an important role in advancing psychiatric research beyond categorical syndromes, transforming our understanding and treatment of mental illness globally.

## Data Availability

NA

## Notes

**Funding Statement –** Research reported here was supported by R01 MH123157 (to LMOL, CLJ, and NBF), R01 MH113078 (to CEB, CLJ, and NBF), R00 MH116115 (to LMOL), T32 MH073526 (to JFDLH) and the Fulbright Commission in Colombia under the Fulbright-Colciencias grant (to JFDLH). The content is solely the responsibility of the authors and does not represent the official views of the Fulbright Program or the National Institutes of Health.

### Competing Interest Statement

The authors have declared no competing interest.

### Funding Statement

Research reported here was supported by R01 MH123157 (to LMOL, CLJ, and NBF), R01 MH113078 (to CEB, CLJ, and NBF), R00 MH116115 (to LMOL), T32 MH073526 (to JFDLH) and the Fulbright Commission in Colombia under the Fulbright-Colciencias grant (to JFDLH). The content is solely the responsibility of the authors and does not represent the official views of the Fulbright Program or the National Institutes of Health.

### Author Declarations

This study was approved by the Institutional Review Board of the University of California, Los Angeles, the Comite de Etica del Instituto de Investigaciones Medicas at Universidad de Antioquia, and the Comite de Bioetica at the Clinica San Juan de Dios Manizales.

### Summary of Updates

This version is more succinct in the introduction and discussion. Some methodological details were moved to the supplementary materials. We added an estimation of the time it takes to reach diagnostic stability in SMI.

