## Supplementary Notes for "Transdiagnostic Clinical Features Delineate Trajectories of Serious Mental Illness"

#### Supplementary Note 1. Primary diagnosis classification and reliability estimation

In a subsample of SMI patients, we assessed the reliability of the ICD-10 diagnoses recorded in the EHR in comparison with those made by an expert research clinician (MC) performing a complete manual chart review. To enable a sufficiently precise estimation of the degree of agreement between these two sets of diagnoses, we selected 120 patients for this record review, chosen at random from among participants whom we had previously recruited at CSJDM in an ongoing study of BD, MDD, and SCZ (12). Of these individuals (40 from each of the three diagnostic groups), we excluded 15 whose most recently recorded ICD-10 diagnoses (F318, F319, F321, F328, or F339) were not among the codes that met our criteria for SMI, as defined above. The clinician review of the remaining 105 records yielded the assignment of a current primary diagnosis based on DSM-5 criteria, as well as a checklist of symptoms and other clinical features of SMI (see below). In these 105 patients, we evaluated (using Cohen’s kappa statistic (14)) the agreement between the diagnosis from the chart review and the most recently recorded ICD-10 SMI code. The agreement between these diagnoses was at a level typically considered “very good” to “excellent” for such comparisons (19,20), with kappa estimates for specific diagnoses considering both inpatient visits and outpatient visits (Supplementary Table 3) were 0.74 (95% CI: 0.60-0.89) for MDD, 0.74 (95% CI: 0.60-0.87) for BD, 0.90 (95% CI: 0.81-0.99) for SCZ; overall kappa=0.78 (95% CI: 0.69-0.88). Positive predictive values (PPVs) were 0.84 for MDD, 0.80 for BD, and 0.92 for SCZ. Estimates for kappas and PPVs were similarly high when considering inpatient visits only (Supplementary Table 3). These levels of agreement are also similar to those from previous studies comparing diagnoses from ICD codes recorded in the EHR with diagnoses from manual chart reviews (21).

#### Supplementary Note 2. NLP algorithm to extract clinical features

The identification of specific clinical features at scale in clinical notes requires an NLP algorithm (15). We, therefore, developed a Spanish-language NLP algorithm to extract features from SMI patients in the CSJDM database. Here, we describe the procedures used to develop, train, and validate this algorithm.

***Overview:*** two clinicians independently reviewed a randomly selected sample consisting of 3,600 passages of free-text (which we term “sentences”) from the inpatient notes, progress notes, and outpatient notes of 2,788 unique SMI patients. During review, they flagged those sentences in which any of the four features were present. 3,310 of these sentences were used to develop the algorithm, and 290 were doubly annotated (i.e., gold standard) and used to evaluate the algorithm’s performance. For the second round of annotation, each putative feature in the evaluation set was labeled as affirmed or negated. Our algorithm involves two distinct tasks: named entity recognition (NER) and negation detection (ND). NER involves identifying instances of a particular clinical feature in the text, for which we utilized the EntityRuler component of the spaCy NLP library (v2.4). ND assesses whether a feature is affirmed or negated within a sentence. To perform this task, we implemented the NegEx algorithm (Chapman et al. 2001). To designate a clinical feature as being “present” for a given patient, we required affirmative mentions of the feature in at least two notes in a patient’s EHR. We evaluated this feature extraction against the checklist compiled by the clinician from the chart review described in the section above. Finally, as an evaluation of the attributions made by the clinician conducting the manual review, we conducted an additional set of manual reviews of selected records; two additional clinicians conducted independent reviews of each of the charts with false positive instances (and an equal number of randomly chosen charts with true positives).

***Sampling of sentences:*** We focused on extracting four specific clinical features: Suicide Attempt, Suicidal Ideation, Delusions, and Hallucinations. In the EHR, we identified nine sections most likely to contain these features. These sections span across three types of notes. **Intake note:** (1) chief complaint, (2) thought content, (3) sensory perception, and (4) assessment. **Progress note**: (5) subjective, (6) objective, and (7) assessment. **Outpatient note**: (8) subjective and (9) assessment.

Notes are represented in the EHR database in the form of tables, where each type of note is stored in its corresponding table. In each of these three tables, a row represents an individual note taken at a specific time for a specific patient (i.e., the index is patient ID and time), while the columns represent distinct sections of a note. For example, in the table for Intake Notes, a row may be the first note of patient X and the columns will be the chief complaint, the thought content, and so on. Each cell in the table, then, contains brief text describing one section of a patient’s note. These text may be as small as a single sentence. Additional columns in the table contain other relevant information for the note, such as the primary ICD-10 code associated with each note (i.e., with each row). The rows in each table were selected to keep only those associated with an ICD-10 code of an SMI diagnosis (F20X, F22X, F25X, F301, F302, F310, F311, F312, F313, F314, F315, F316, F317, F322, F323, F331, F332, F333, or F334), resulting in 19,713 intake notes, 225,362 progress notes, and 26,673 outpatient notes.

From these filtered tables, we isolated each of the nine columns that represent the sections identified above. Finally, from each column we randomly sampled 400 cells for annotation, i.e., a total of 3600 cells across the nine columns. The text contained in each of the 3600 cells is hereafter referred to as a “sentence”.

***Annotation of clinical features:*** Two clinicians independently annotated each sentence for the presence of the four clinical features, identifying the specific span of text inside the sentence in which each feature was mentioned. For example, in the sentence, “The patient is presenting with auditory hallucinations,” the text “auditory hallucinations” would be identified, and the sentence flagged for the presence of the feature – Hallucinations.

***Gold Standard and Development Set Creation:*** Of the 3600 sentences, at least one annotator flagged 83 for Suicide Attempt, 523 for Suicidal Ideation, 317 for Delusions, and 495 for Hallucinations. We selected at random 30% of each one of these four sets of sentences to generate a gold standard. The sentences in these sets may overlap with each other, i.e., sentences selected based on one feature may contain additional features; for example, “the patient is presenting with persecutory *delusions* and *suicidal ideation*”.

In total, 290 sentences were included in the gold standard; in these sentences all inconsistencies between annotators were resolved, and each clinical feature was labeled as either affirmed or negated. The remaining 3310 sentences were used as the development set. For each of the four clinical features in the development set, we counted how many sentences were flagged by either annotator or by both, and estimated the inter-annotator agreement using Cohen’s kappa (Cohen 1960). These kappas indicated “good” to “excellent” agreement between two independent annotators for all four features (Supplementary Table 4).

***Algorithm for Clinical Feature Extraction*:** The first step was to develop the list of search patterns to be used in NER. For this, we used the spans of text identified inside the sentences of the development set. Concretely, these spans of text are sequences of words that represent a clinical feature. We converted each span to lowercase and formatted it using two different components of spaCy’s medium-sized Spanish language model (sp_news_md), as follows. First, we used the “tokenizer” component to split the span of text into a sequence of words and punctuation marks (jointly known as tokens). Then, we used the “tagger” component to assign a part-of-speech (POS) tag to each token. Within each sequence of tokens, we replaced the tokens with their POS-tags, unless their tags were one of the following: noun, verb, adjective, adverb, pronoun, auxiliary or subordinating conjunction. In such cases, the tokens were replaced by their lemma. As a result, a search pattern is a sequence of POS-tags and lemmas of the same length and order as the sequence of tokens identified during annotation. (Supplementary Table 5). The final list of search patterns was manually curated to increase the coherence of the clinical feature. The procedures for this manual curation included: first, removing patterns that, by themselves, were not sufficiently complete to ensure that they indicated the presence of the feature (e.g., the pattern “visual” is not sufficient to indicate Hallucinations); then, delineating the boundaries of the features by excluding patterns that could not be unambiguously interpreted as indicating the presence of the feature (e.g., separating Suicide Attempt from self-harm). These procedures reduced the initial number of patterns for each feature, from 44 to 23 in Suicide Attempt, from 176 to 78 in Suicidal Ideation, from 127 to 119 in Delusions and from 154 to 122 in Hallucinations. Finally, the curated list of patterns was passed to the “EntityRuler” component of spaCy to complete the NER task.

Subsequently, we detected the negation status of each clinical feature using the NegEx algorithm. Briefly, this algorithm assumes each feature to be “affirmed” by default and its status is only changed to “negated” when it is located within five tokens of a negation cue. Negation cues were manually identified in the development set (Supplementary Table 6) and their location in the text was determined using the “EntityRuler” component of spaCy.

To improve the accuracy of both patterns and negation terms, we ran our full pipeline (NER and ND) with each of the four features, on the entire EHR database and randomly selected 100 instances (50 affirmed and 50 negated) of each feature for manual review. Two clinicians evaluated the 400 instances and recommended adjustments to the search patterns and negation cues. Although this process can be iterated until the expected performance is achieved, we considered a single iteration to be sufficient for all features.

***Validation of extracted features*:** To evaluate the performance of our algorithm for each one of the four clinical features, we report precision, recall, and F1 (i.e.: the harmonic mean of precision and recall). First, at the level of individual sentences, we used the 290 sentences of the gold standard described above. Our algorithm performed with a high rate of precision (range: 0.88-1.0) and recall (range: 0.62-1.0), resulting in a satisfactory F1-score for all features (suicide attempt: 0.82, suicidal ideation: 0.73, delusions: 1.0, hallucinations: 0.95), (Supplementary Table 7, see Supplementary Note 7 for a description of errors). At the patient level, we used the item checklist from the clinician’s chart review of 120 patients described in the main text. For any given patient, a “lifetime” phenotype extracted by our NLP pipeline was defined as follows: by default, the phenotype is absent, and it is changed to present only if the patient has two or more notes with affirmed instances of the clinical feature. One patient out of the 120 had to be excluded from this evaluation because they had only one note on record.

***Threshold for a “lifetime” phenotype from NLP extracted features:*** The threshold of “two notes or more” is arbitrary. We hypothesized that requiring more affirmative mentions of a feature to classify a patient would result in increased precision of the phenotype, while at the same time reducing its recall; that is, increasing the two-note threshold could result in a narrower and likely more severe phenotype, albeit with a smaller sample size. We tested this hypothesis by varying the number of notes required to change a patient’s “lifetime” phenotype from absent to present from 1 to 10 and determined the optimal balance of precision and recall using an F1-score. (Supplementary Figure 2). requiring two such mentions provided an optimal balance (i.e., resulted in maximal F1) between precision and recall. At this threshold, the algorithm output and the designation of features from manual chart review were highly concordant for all four features; suicide attempt (92/104, F1 = 0.68), suicidal ideation (89/104, F1 = 0.79), delusions (84/104, F1 = 0.82), and hallucinations (87/104, F1 = 0.84), (Supplementary Table 7B).

***Addressing human error in chart review:*** Some degree of human error can be expected when performing clinical chart review. Specifically, features may be overlooked in reviewing an extensive clinical history. We therefore conducted an additional manual chart review (by two independent clinicians blinded to the patients’ original classification) of the 29 instances of apparent discordance between the algorithm output (which reported one or more features) and the initial manual review (which had not reported these features). In 16 of these instances both of the subsequent reviewers reported features that the initial reviewer had apparently overlooked. In four additional instances the output was ambiguous, meaning that the two raters disagreed on whether the concept was present. The remaining nine instances were false positives. Agnostic to the observed concordance/discordance of each instance, reviewers also independently examined 29 randomly selected instances for which the algorithm output and the initial review were concordant (both interview and review reported a feature); their reports confirmed the concordance for 28 of these instances, with the last one being ambiguous. We then incorporated the information obtained from this second review in a *post hoc* analysis comparing the algorithm output to manual review, requiring agreement between both raters; we observed an increase in concordance and F1 for all four clinical features (Suicide Attempt: 94/104, F1 = 0.75; Suicidal Ideation: 95/104, F1 = 0.89; Delusions: 88/104, F1 = 0.86 and Hallucinations: 91/104, F1 = 0.88), Supplementary Table 7C).

#### Supplementary Note 3. Patient-level data validation: NLP features and ICD-10 diagnoses

We evaluated the relationship between the presence of *lifetime* clinical features with gender and with the most recent diagnoses of MDD, BD, and SCZ, at the *individual level*.

As established above in our validation of NLP clinical features using chart review, we defined “lifetime” features for patients with at least two notes (n=20,658; Supplementary Figure 1C). We define a feature as present in a patient if at least two notes in their records have an affirmative mention of the feature.

To test for associations between diagnosis, gender, and features, we used logistic regression to model the logit of the probability of a feature being present as a function of gender (female=1), and current diagnosis (BD as reference).

In this model, we accounted for the number of notes a patient has on record using the log_10_ transformed variable $N notes$, since the likelihood of a feature being present is expected to increase with the number of notes. We also account for illness severity by including a binary variable that is 1 if the individual has had a history of hospitalization (*Inpatient*).

Consider the probability that feature *s* is present to be $P_{{Sx}_{s}}$. The resulting model is:

$$ln\left( \frac{P_{{Sx}_{s}}}{1-P_{{Sx}_{s}}} \right) \sim\beta_{0}+\beta_{1}MDD+\beta_{2}SCZ+\beta_{3}Gender+ \beta_{4}{log}_{10}\left( N notes \right)+\beta_{5}Inpatient$$

We fit four models in total, one for each feature. In each model we tested for associations with gender and with two diagnoses (MDD and SCZ), for a Bonferroni-corrected alpha of 0.05/12=0.0041.

Then, to explore the interactions between gender and diagnosis, we expanded the model to include interaction terms. This model is expressed as:

$$ln\left( \frac{P_{{Sx}_{s}}}{1-P_{{Sx}_{s}}} \right) \sim\beta_{0}+\beta_{1}MDD+\beta_{2}SCZ+\beta_{3}Gender+ \beta_{4}{log}_{10}\left( N notes \right)+\beta_{5}Inpatient$$

$$+ \beta_{6}Gender:MDD+\beta_{7}Gender:SCZ$$

To evaluate the relationship between different clinical features that may occur in a patient over the entire course of their EHR, we used the above modelling framework, but added as predictors, for each feature ${Sx}_{s}$, the lifetime presence of the three remaining features ( ${Sx}_{j} ; j\neq s ; s,j\in\left\{ 1,2,3,4 \right\}$ ). The resulting model is:

$$ln\left( \frac{P_{{Sx}_{s}}}{1-P_{{Sx}_{s}}} \right) \sim\beta_{0}+\beta_{1}MDD+\beta_{2}SCZ+\beta_{3}Gender+ \beta_{4}{log}_{10}\left( N notes \right)+\beta_{5}Inpatient$$

$$+\sum_{j=1}^{4} \gamma_{j\neq s}{Sx}_{j\neq s}$$

We fit four models in total, one for each feature. In each model, we tested for associations with gender and with the three remaining features (e.g., in the case of delusions, we tested for associations with suicide attempt, suicidal ideation, and hallucinations). This procedure results in a Bonferroni-corrected alpha of 0.05/16=0.0031.

Finally, to explore the interactions between gender and co-occurring features, we expanded the model to include an interaction term, as follows:

$$ln\left( \frac{P_{{Sx}_{s}}}{1-P_{{Sx}_{s}}} \right) \sim\beta_{0}+\beta_{1}MDD+\beta_{2}SCZ+\beta_{3}Gender+ \beta_{4}{log}_{10}\left( N notes \right)+\beta_{5}Inpatient$$

$+\sum_{j=1}^{4} \gamma_{j\neq s}{Sx}_{j\neq s}+\sum_{j=1}^{4} \alpha_{j\neq s}{Gender:Sx}_{j\neq s}$

#### Supplementary Note 4. Definition of diagnostic trajectories and examples

To define a diagnostic trajectory, we map the progression of a patient's diagnoses over time using the sequence of ICD-10 codes extracted from their EHR. Starting with a patient's initial diagnosis, each subsequent visit contributes to this trajectory by either introducing a new diagnosis or indicating a switch from a previous diagnosis. Specifically, we followed these steps:

1. Record the diagnosis from the patient's first visit.
2. At every subsequent visit, add the new diagnosis to the patient’s cumulative record unless the diagnosis is already there.
3. If the diagnosis is found to be incompatible with a pre-existing one (as determined in Supplementary Table 2), the prior diagnosis is replaced, marking a diagnostic switch.

In the resulting trajectory, consecutive visits that do not introduce new diagnoses are condensed to avoid redundancy, ensuring that the final trajectory primarily represents either the acquisition of new comorbidities or diagnostic switches. This approach offers a concise and chronological representation of a patient's diagnostic journey over time. We provide below, two working examples of this procedure:

Consider the following sequence of ICD-10 diagnoses in the EHR of a patient with five visits.

**F32 -> F32 -> F31 -> F31 -> F31**

Here, the patient’s initial diagnosis of F32 (MDD) switched to F31 (BD) by the third visit. This trajectory incorporating the switch can be represented by:

**F32 -> F31**

Consider now the following sequence of ICD-10 diagnoses in the EHR of a patient with six visits.

**F32 -> F41 -> F32 -> F32 -> F31 -> F31**

Here, the patient had an initial diagnosis of F32 (MDD). On their second visit, they acquired the comorbidity of F41 (anxiety disorders) and finally the diagnosis switched from F32 to F31 (BD) on the fifth visit. This trajectory incorporating both comorbidity and a switch can be represented by:

**F32 -> F32,F41 -> F31,F41**

#### Supplementary Note 5. Factors contributing to visit-to-visit diagnostic stability

We used visit-level data to characterize the rate at which diagnoses stabilize over time and the factors that increase or decrease diagnostic stability.

To do this, we modeled a switch in diagnosis using the binary variable ${Switch}_{k+1}$. A value of 1 indicates that a patient’s diagnosis at their next visit (*k+1)* is different from their current one:

$${Switch}_{k+1}: {ICD10}_{k}^{Dx} \neq{ICD10}_{k+1}^{Dx}$$

We fit a mixed-effect logistic regression with the logit of the probability of a diagnostic switch in visit k+1 as the outcome and (log-transformed) visit number *k* as the predictor. We accounted for repeated observations of patients using random intercepts. We define $P_{{Switch}_{k+1,i}}$ as the probability of a diagnostic switch in visit k+1 for patient i. The resulting model is:

$$ln\left( \frac{P_{{Switch}_{k+1,i}}}{1-P_{{Switch}_{k+1,i}}} \right) \sim\beta_{0}+\beta_{1}{log}_{10}\left( k_{i} \right)+u_{0i}+e_{i}$$

Where $u_{0i}\sim N\left( 0,{\sigma_{u}}^{2} \right)$ is a random intercept with mean 0 and variance ${\sigma_{u}}^{2}$ and $e_{i}\sim N\left( 0,{\sigma_{e}}^{2} \right)$ is residual error. In this framework, if a patient has *K* total visits, they contribute *K – 1* observations to the analysis, since their final visit doesn’t have a ${Switch}_{k+1}$.

We used this flexible framework to understand the short-term stability of diagnoses. For this, we included dummy variables to indicate the ICD-10 diagnosis at visit k, with BD as the reference. Possible diagnoses were *MDD, SCZ, other*. We extended this model with four additional explanatory variables, two at the visit level and two at the patient level. At the visit level these explanatory variables are represented by two binary indicators: inpatient status (${Inpatient}_{k}$) and an indicator representing an ICD-10 diagnosis of “Not Otherwise Specified” (NOS; ${ICD10}_{k}^{NOS}$). These were defined as all codes with the form FXX8 ("Other ...") or FXX9 ("Unspecified ..."), as well as those with the form FX8 and FX9 that are explicitly named "Other [...] Disorder" or "Unspecified [...] Disorder", respectively. At the patient level, we included gender and age at first visit. Additionally, we included a binary variable for each of the four clinical features (${Sx}_{j,k}$, see Supplementary Note 2), indicating if they were present during the current visit.

The resulting model is:

$$ln\left( \frac{P_{{Switch}_{k+1,i}}}{1-P_{{Switch}_{k+1,i}}} \right) \sim\beta_{0}+\beta_{1}{log}_{10}\left( k_{i} \right)+\beta_{2}{MDD}_{k,i}+\beta_{3}{SCZ}_{k,i}+\beta_{4}{other}_{k,i}+\beta_{5}{Inpatient}_{k,i}+\beta_{6}{ICD10}_{k,i}^{NOS}$$

$$+ \beta_{7}{Gender}_{i}+\beta_{8}{Age}_{i}+\sum_{j=1}^{4} \gamma_{j\neq s}{Sx}_{j\neq s,k,i}+u_{0i}+e_{i}$$

Finally, to assess the evidence of sustained diagnostic instability, we fit a second model where we include the switch indicator for the previous visit – ${Switch}_{k}$ – to estimate the effect of a previous switch on a future switch. In this analysis, each patient contributes *K– 2* observations to the analysis, since their first visit doesn’t have a ${Switch}_{k}$. This model is:

$$ln\left( \frac{P_{{Switch}_{k+1,i}}}{1-P_{{Switch}_{k+1,i}}} \right) \sim\beta_{0}+\beta_{1}{log}_{10}\left( k_{i} \right)+ \beta_{2}{MDD}_{k,i}+\beta_{3}{SCZ}_{k,i}+\beta_{4}{other}_{k,i}+\beta_{5}{Inpatient}_{k,i}+\beta_{6}{ICD10}_{k,i}^{NOS}$$

$$+ \beta_{7}{Gender}_{i}+\beta_{8}{Age}_{i}+\beta_{9}{Switch}_{k,i}+\sum_{j=1}^{4} \gamma_{j\neq s}{Sx}_{j\neq s,k,i}+u_{0i}+e_{i}$$

For this section, we use a Bonferroni-corrected alpha of 0.05/13=0.0038.

#### Supplementary Note 6. Prediction of diagnostic codes from NLP-extracted clinical features:

We explored if NLP-extracted features can forecast future diagnostic codes. Specifically, we assessed if clinical features extracted from free-text during a patient's current visit (k) can predict ICD-10 codes for the subsequent visit (k+1). For major mood disorders (MDD and BD), the 3-digit ICD codes at each visit indicate the episode's severity and the presence or absence of psychotic features (Supplementary Figure 1E). Within visits with codes for severe episodes, we represented episodes with and without psychotic symptoms with the binary variable ${ICD10}_{k}^{psychosis}$. This variable is 1 for codes: F302, F312, F315, F323, or F333. And 0 with codes: F301, F311, F314, or F322. We also defined the clinical feature, “Psychosis” (${Sx}_{p}$), to include the presence of either Delusions or Hallucinations extracted from the notes.

Using a logistic regression, we evaluated whether the presence of this feature during a visit *k* is associated with the logit of the probability of an ICD-10 code indicative of an episode *with* psychotic symptoms in the following visit *k+1*. This clinical feature is more frequently found in the notes from inpatient visits than in notes from outpatient or emergency department visits; this observation likely reflects not only the increased severity of symptoms experienced by individuals in association with inpatient hospitalization, but also the larger number of notes recorded during an inpatient stay. To account for the latter factor, we adjust for the binary variable ${Inpatient}_{k}$.

To fit this model, we adjusted for the presence of psychosis in visit k (${ICD10}_{k}^{psychosis}$, defined as above), the clinical feature ${Sx}_{p}$ on visit k+1 (${Sx}_{p,k+1}$), and the inpatient status in both visits (${Inpatient}_{k}$ and ${Inpatient}_{k+1}$). Let $P_{{ICD10}_{k+1,i}^{psychosis}}$ be the probability that patient *i* has a psychosis ICD-10 code at visit *k+1*. Finally, we account for repeated observations on a patient with a random intercept as above. The resulting model is:

$$ln\left( \frac{P_{{ICD10}_{k+1,i}^{psychosis}}}{1-P_{{ICD10}_{k+1,i}^{psychosis}}} \right) \sim\beta_{0}+\beta_{1}{Sx}_{p,k,i}$$

$$+ \beta_{2}{ICD10}_{k,i}^{psychosis}+\beta_{3}{Sx}_{p,k+1,i}+\beta_{4}{Inpatient}_{k,i}+\beta_{5}{Inpatient}_{k+1,i}+u_{0i}+e_{i}$$

Where $u_{0i}\sim N\left( 0,{\sigma_{u}}^{2} \right)$ is a random intercept with mean 0 and variance ${\sigma_{u}}^{2}$ and $e_{i}\sim N\left( 0,{\sigma_{e}}^{2} \right)$ is residual error.

#### Supplementary Note 7. Evaluation of errors in the gold standard.

Considering both false positives and false negatives, the NLP algorithm failed in 28 instances of the gold standard. These errors happened in both the NER step and in the ND step.

Most errors occurred in the NER step (25/28) and were due to missing search patterns. We identified three reasons for missing patterns: 1) the text contains spelling errors that were not observed in the development set; 2) the specific pattern did not appear in the development set; and 3) the pattern was observed in the development set but was removed because it was not specific enough and would have generated numerous false positives.

Three false positives were caused by failure to identify the negation in the sentence. In two of these instances, the feature was part of a list of negated terms and was located beyond the scope of the negation cue (five tokens). In the last instance, the error was caused by a spelling error in the negation cue that was missing from the development set.
