## Supplementary Figures for "Transdiagnostic Clinical Features Delineate Trajectories of Serious Mental Illness"

#### **Supplementary Figure SF1.** Flow diagram of sample selection from the CSJDM EHR database indicating: (A) the steps used to remove patients and patient visits not meeting criteria for any of our analyses; (B) the complete SMI cohort from which we selected subsets for different analyses as described in the Methods; (C) Cohort for evaluating patient-level association between clinical features and ICD-10 diagnoses (2+ notes); (D) Cohort for the trajectory analyses exploring diagnostic switches and comorbidities (3+ visits) (E) Visits used for testing if clinical features identified at one visit anticipate ICD-10 code changes at the subsequent visit (consecutive visits with a code for severe mood episode); (F) Cohort for estimating time-to-diagnostic stability (10+ years). Cohorts C-F are not mutually exclusive. Patients may appear in multiple cohorts depending on which criteria they meet. For example, a patient with 8 visits over 11 years and 20 notes will be included in cohorts C, D and F.

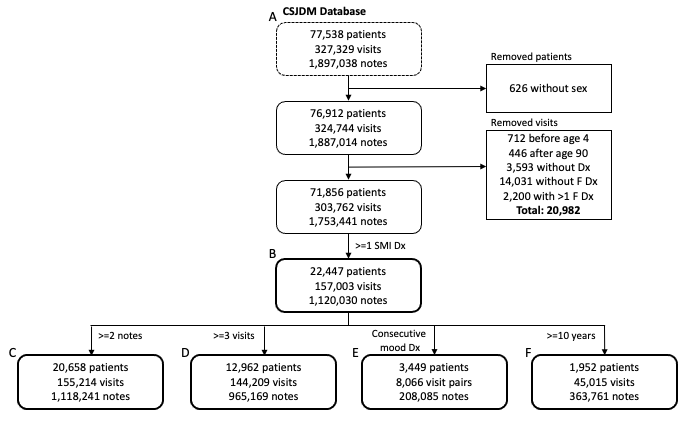

#### **Supplementary Figure SF2.** The performance of the NLP algorithm at different thresholds for the number of affirmative mentions required to classify patients as positives or negatives for each clinical feature. We considered all thresholds between one to ≥10 affirmative mentions per patient; we could only evaluate such mentions if a patient had at least that number of different notes in their EHR, and therefore the sample size of patients who could be evaluated decreases with increasing thresholds (from 105 patients with ≥ one note to 88 with ≥ 10 or more notes). At each threshold we evaluated the performance of the algorithm in terms of precision, recall and F1. We selected a threshold of ≥ two affirmative mentions to designate a patient as positive for a clinical feature, as this threshold (evaluated in the119 patients with at ≥ two notes) yields the highest F1 across the four features.

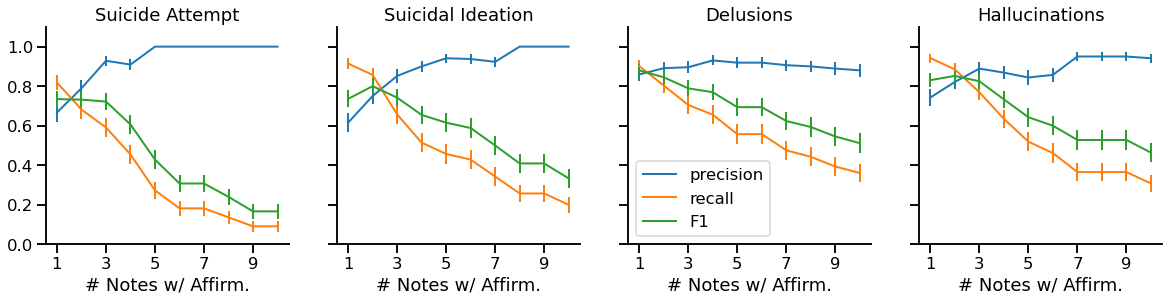

#### **Supplementary Figure SF3.** Sankey diagram of ICD-10 code trajectories. The figure shows switches between SMI diagnoses in patients with 3 or more visits. N=12,962 patients.

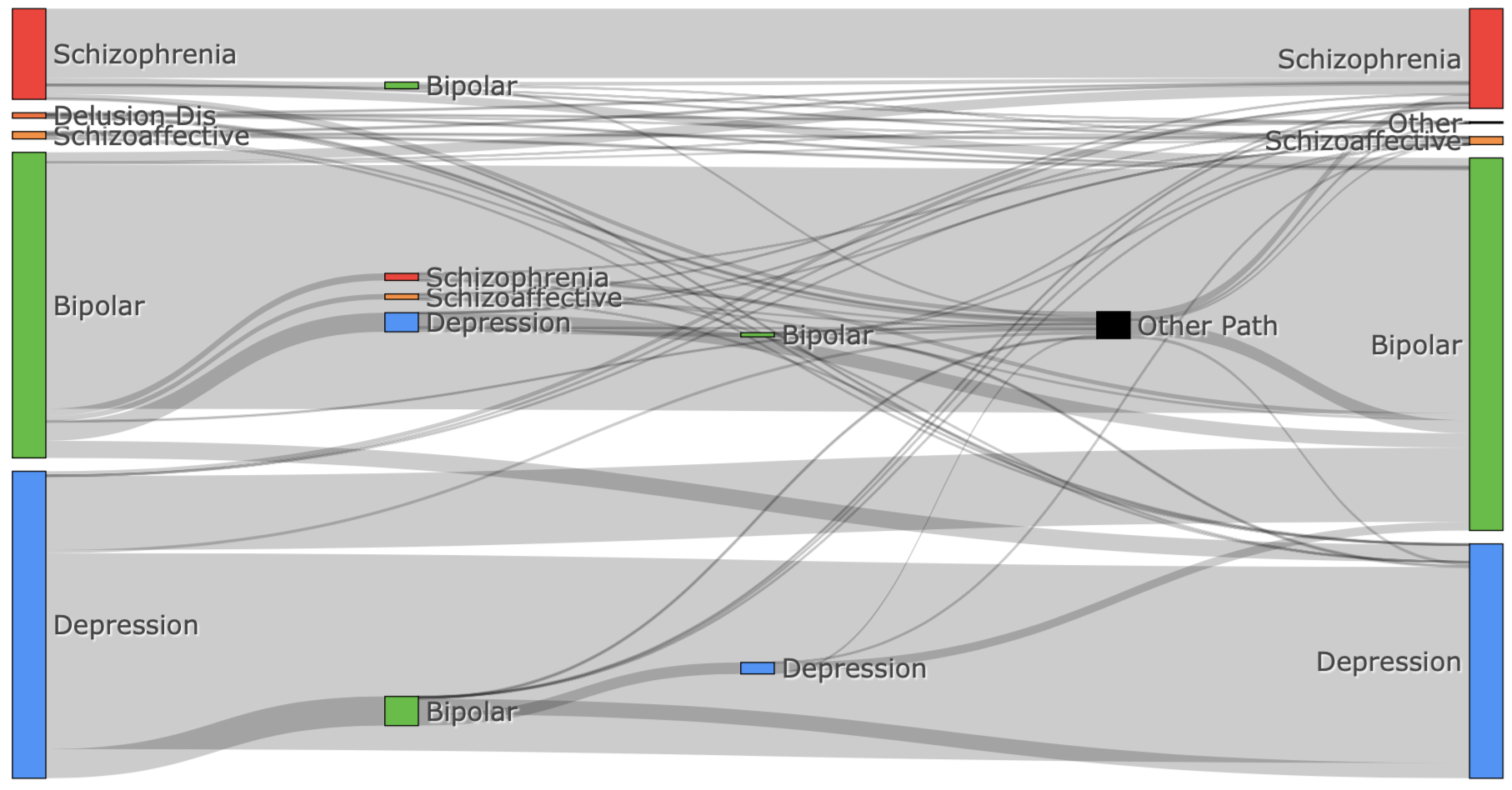

#### **Supplementary Figure SF4.** Distribution of visit number at which specific switches occur. Switches shown occur more than 100 times and are sorted by the average visit at which they occurred. Early switches include brief psychotic disorder (F23) to BD (F31); later switches include SCZ (F20) to schizoaffective disorder (F25). N=12,962 patients.

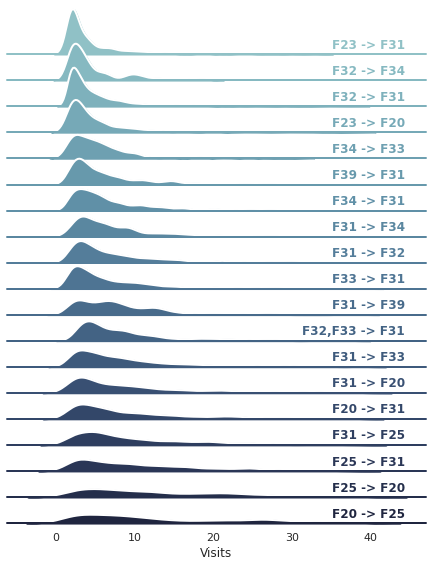

#### **Supplementary Figure SF5.** Future instability as a function of previous instability. The figures show the diagnostic instability (i.e.: the proportion of visits that will have a diagnostic switch on the next visit) over time measured as A) visit number and B) years since the first visit. Proportions are stratified by whether the current visit was a diagnostic switch from the previous visit. The dots correspond to observed probabilities and the solid lines correspond to the probabilities estimated by the model. Whiskers and shared areas correspond to 95% confidence intervals. N=12,962 patients.

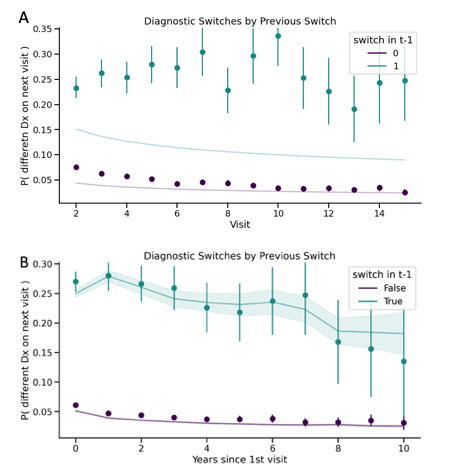
