## Supplementary Tables for "Transdiagnostic Clinical Features Delineate Trajectories of Serious Mental Illness"

#### **Supplementary Table 1.** ICD-10 codes used in this study. Codes meeting our definition of SMI are marked with *.

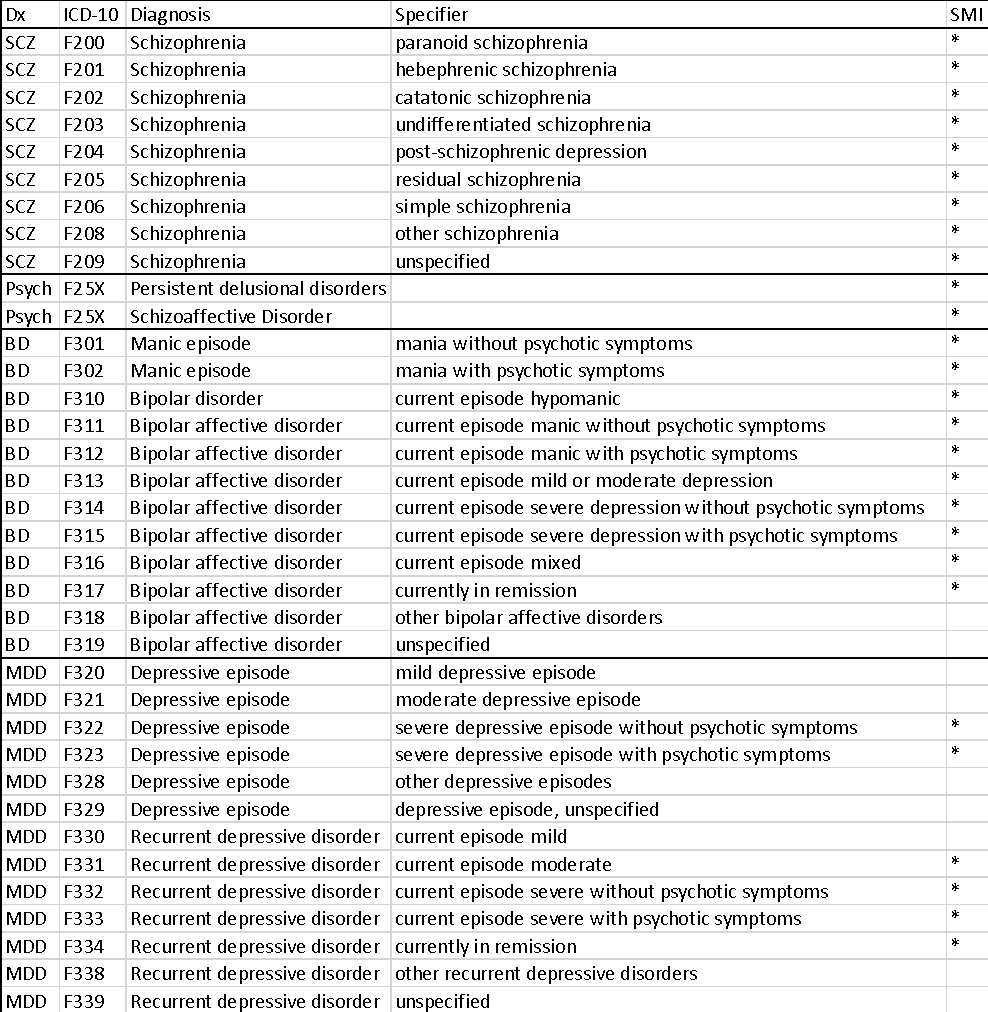

#### **Supplementary Table 2.** List of diagnostic pairs from the F chapter of ICD-10 that are, by definition, incompatible with each other and, therefore, represent diagnostic switches. All other combinations of diagnoses are considered comorbidities. There are two exceptions to this rule: the pairs F30-F31 and F32-F33. These are neither switches nor comorbidities.

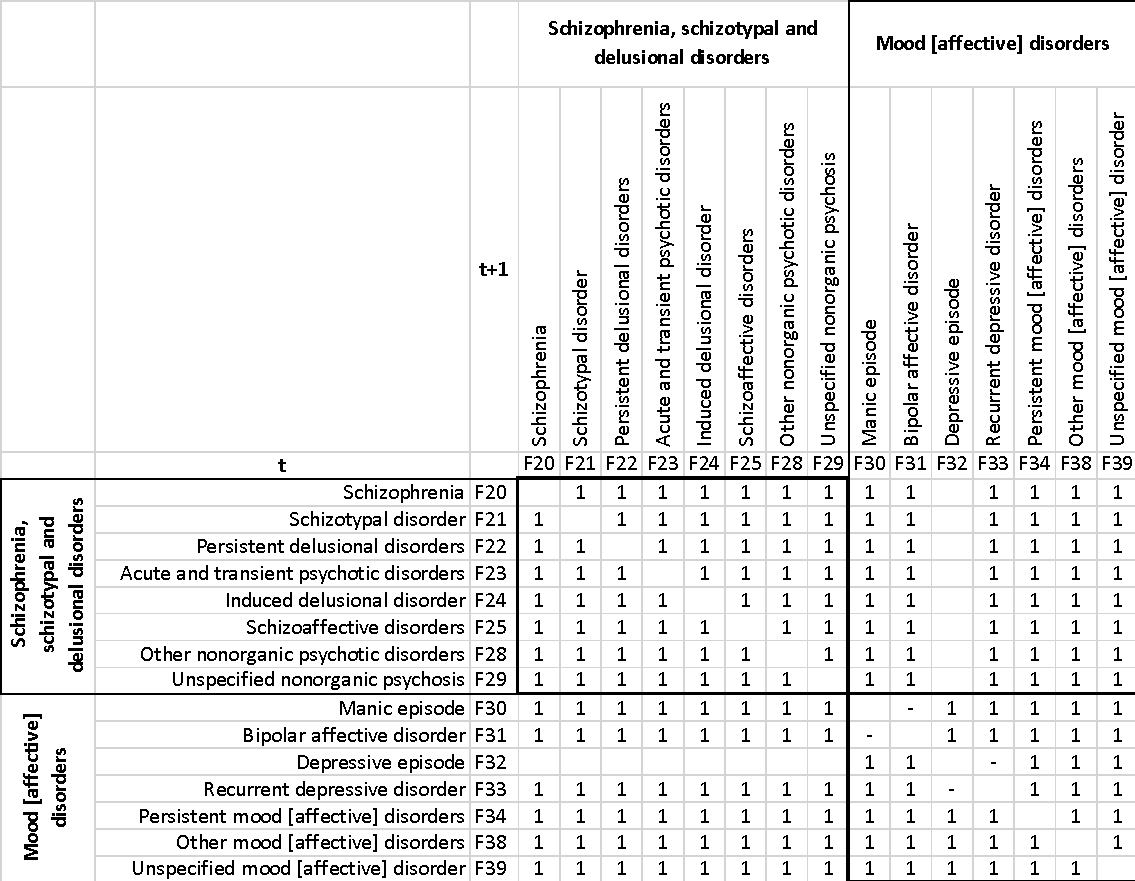

#### **Supplementary Table 3.** Estimates of kappa and PPV from comparisons of ICD-10 diagnoses extracted from the EHR and clinician diagnoses obtained from chart review, considering all visits and inpatient visits only. The narrow definition refers to the SMI codes: F20X (SCZ), F301, F302, F310, F311, F312, F313, F314, F315, F316, F317 (BD), F322, F323, F331, F332, F333, F334 (Severe/Recurrent MDD). The broad definition encompasses, additionally, all F31X (including F318 and F319), F32X (including F320, F321, F328 and F329) and F33X (including F330, F338 and F339). Kappa values are estimated both for individual diagnoses and across all diagnoses. 95% confidence intervals for kappa values are shown in parentheses. Kappa values between 0.6-0.8 are considered “very good”, while those > 0.8 are considered “excellent”.

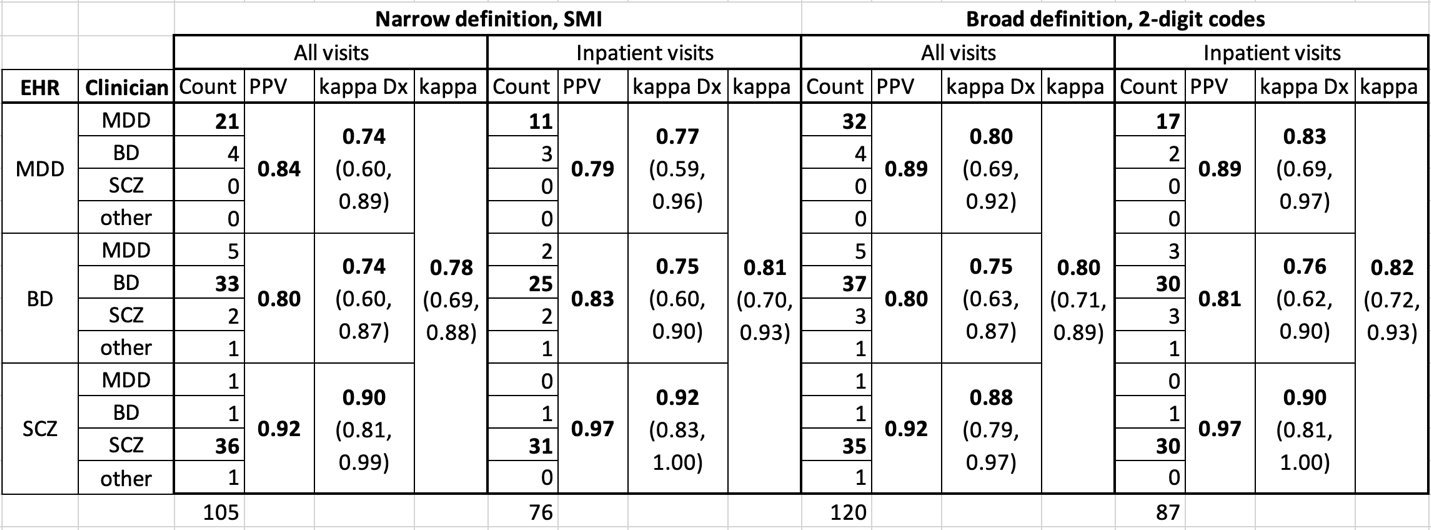

#### **Supplementary Table 4.** Annotation results for four clinical features. Two clinicians independently reviewed and annotated 3,600 sentences. The columns show the number of sentences containing the clinical features identified by Clinician A (Clin. A), Clinician B (Clin. B), or either clinician (Union), and their level of agreement as estimated by Cohen’s kappa (Kappa). Kappa values between 0.4-0.6 are considered “good,” those between 0.6-0.8 are considered “very good,” and those > 0.8 are considered “excellent.”

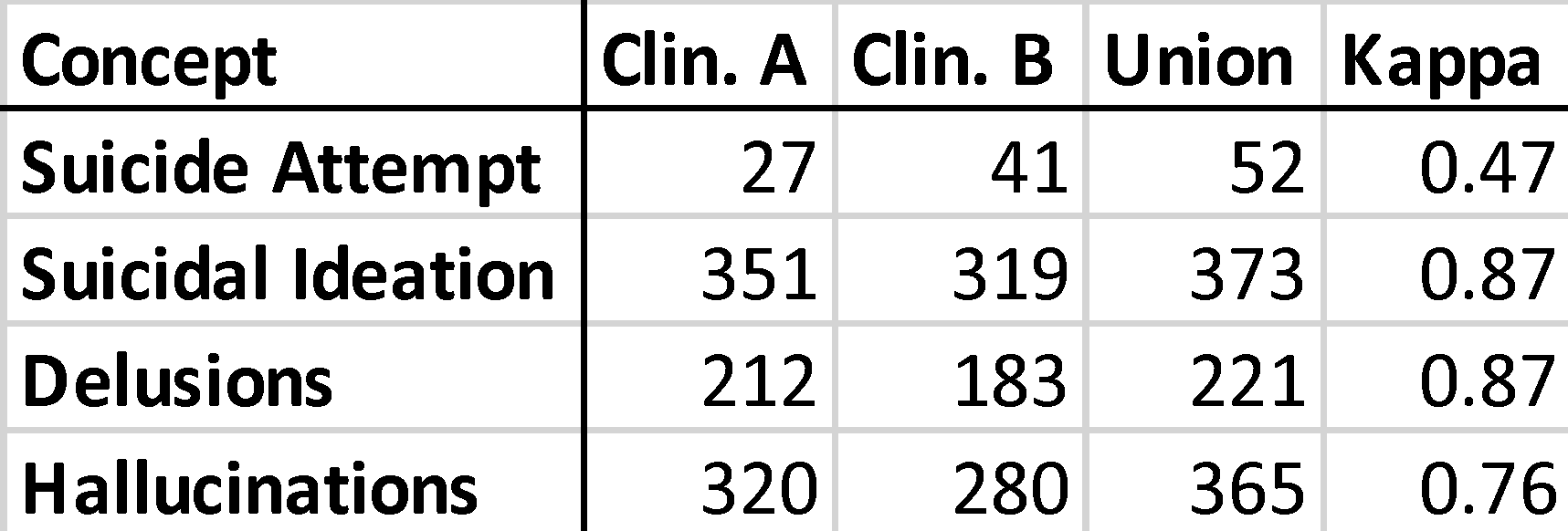

#### **Supplementary Table 5:** Patterns that the NLP algorithm uses for identifying clinical features in the notes. *Label*: label used for annotating clinical features. SUI_ATTP: Suicide Attempt, SUI_IDEA: Suicidal Ideation, DEL: Delusions, HAL: Hallucinations; *Pattern*: sequence of tokens used by the EntityRuler component of Spacy to perform Named Entity Recognition; *Annotation*: specific span of text highlighted by the annotators and used to generate the pattern (as described in Supplementary Note 1); *Notes*: number of notes represented by the same annotation; *Source*: indicates if the pattern derives from the annotation or the subsequent curation process

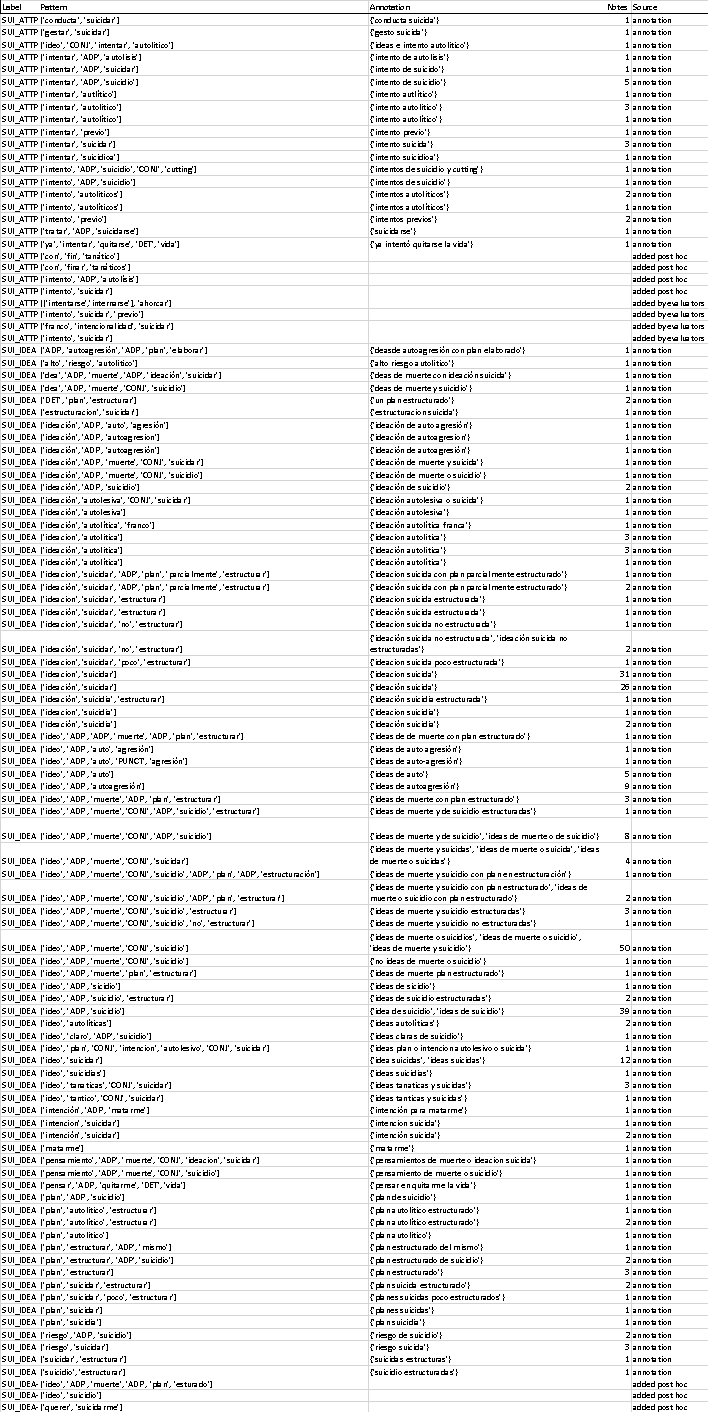

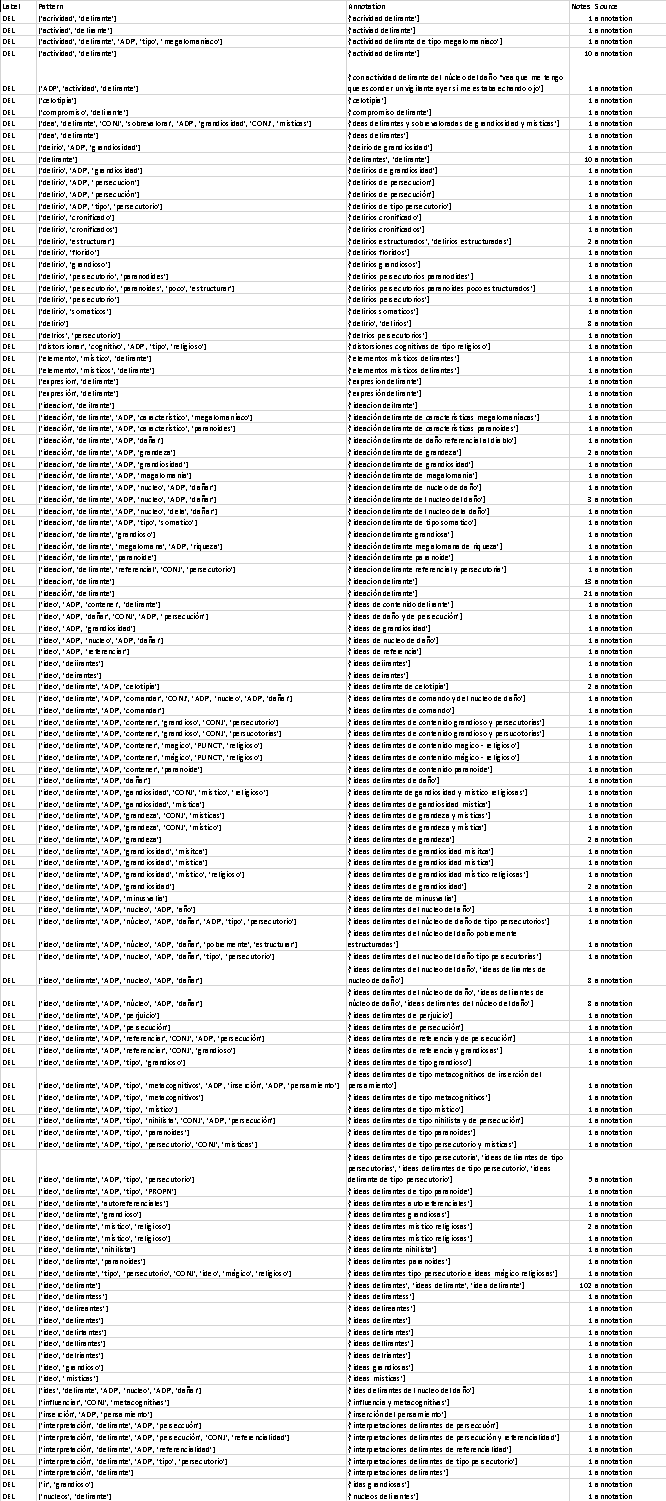

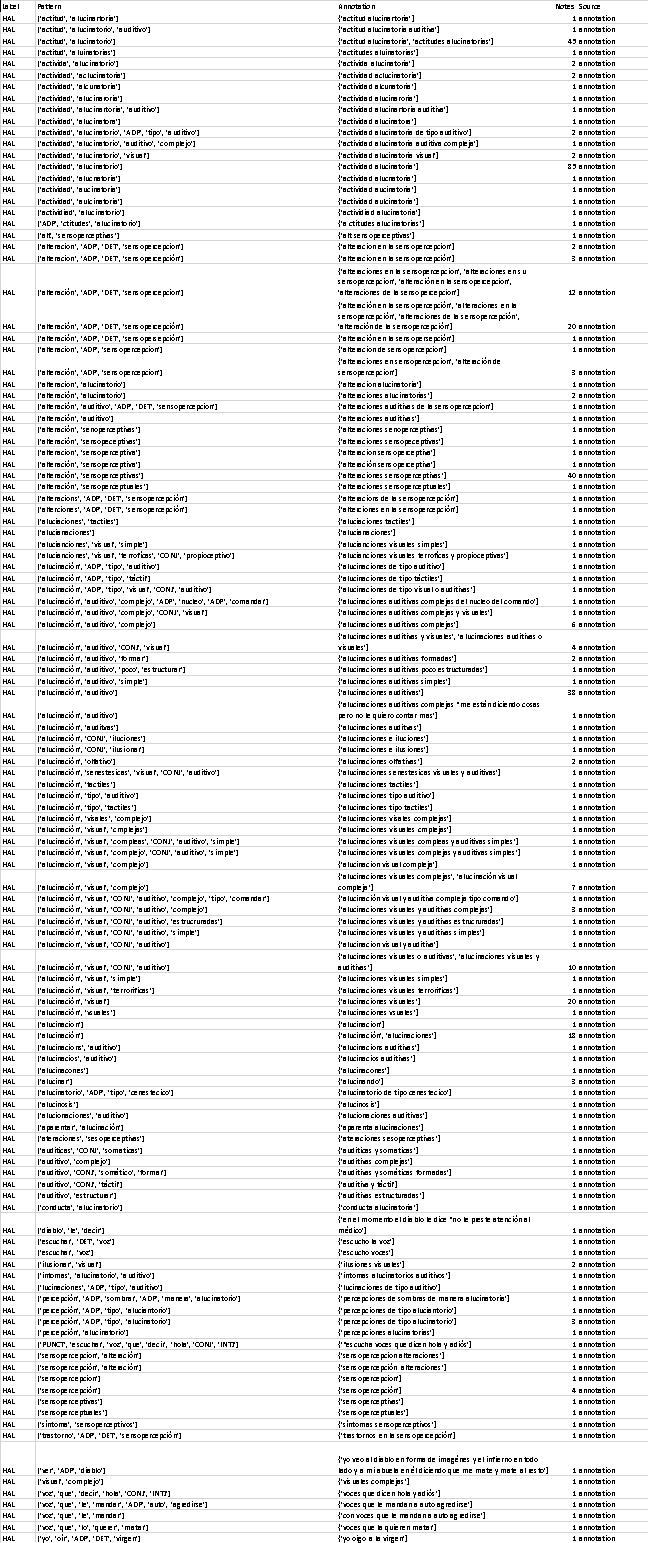

#### **Supplementary Table 6:** Patterns used by the NegEx algorithm. Type: either negation, pseudo-negation, or termination of scope. Scope: directionality of negation.

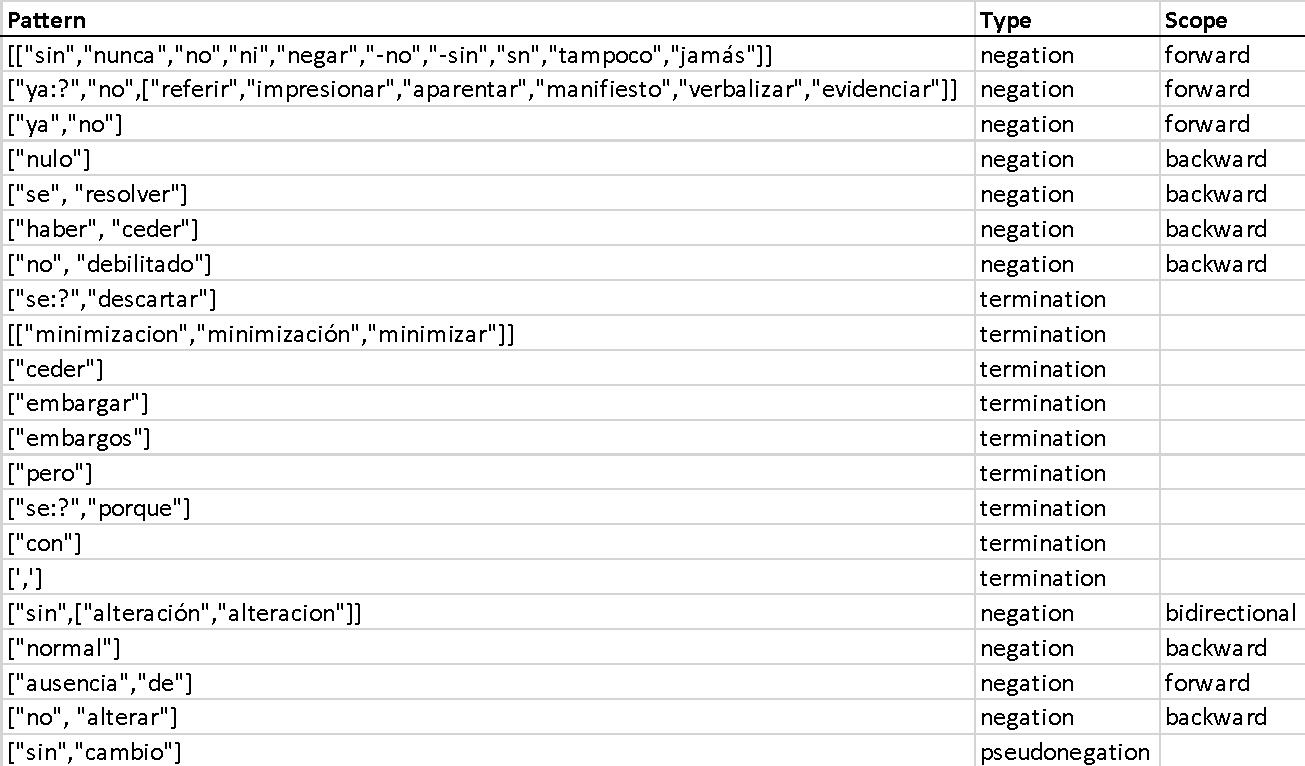

#### **Supplementary Table 7. Performance of the NLP algorithm for extraction of clinical features**. A) Sentence-level performance on the annotated gold standard. B) Patient-level performance on patient records manually reviewed by a clinician (n=104, as one patient was removed for having only one clinical note). C) Patient-level performance after post-hoc review of true and false positives. The average affirmative and negative instances of each feature per patient are, respectively, 1 and 0 for suicide attempt, 4 and 12 for suicidal ideation, 17 and 19 for delusions, and 10 and 25 for hallucinations.

A)
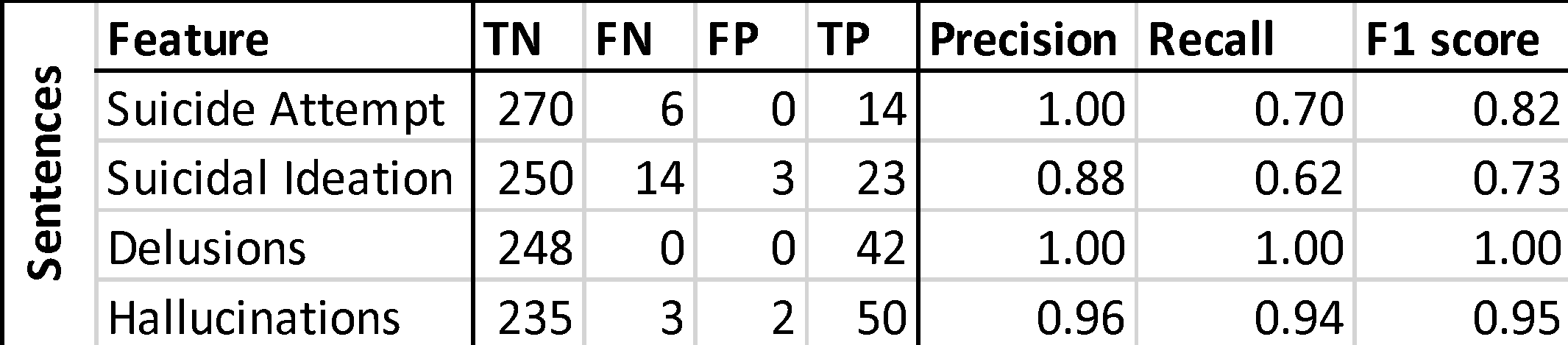

B)
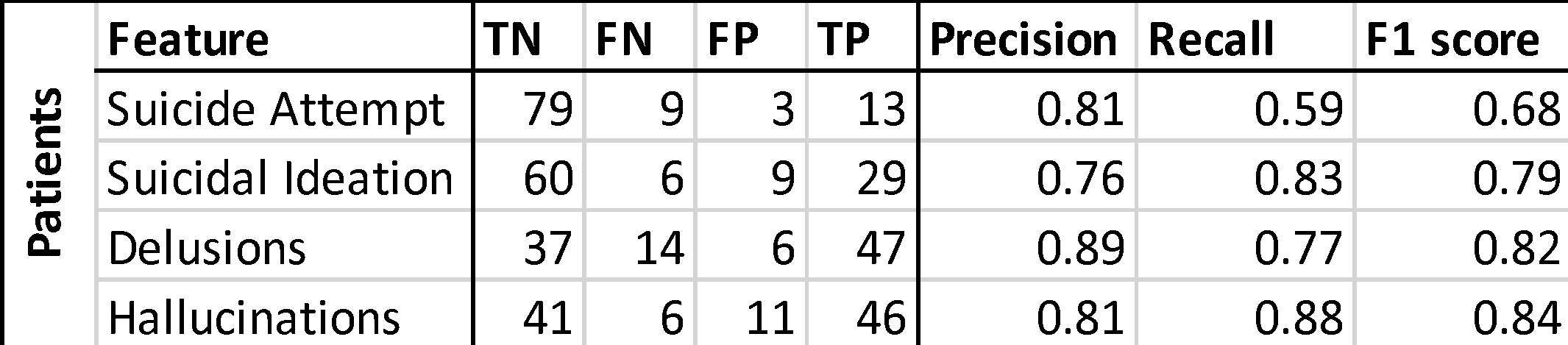

C)
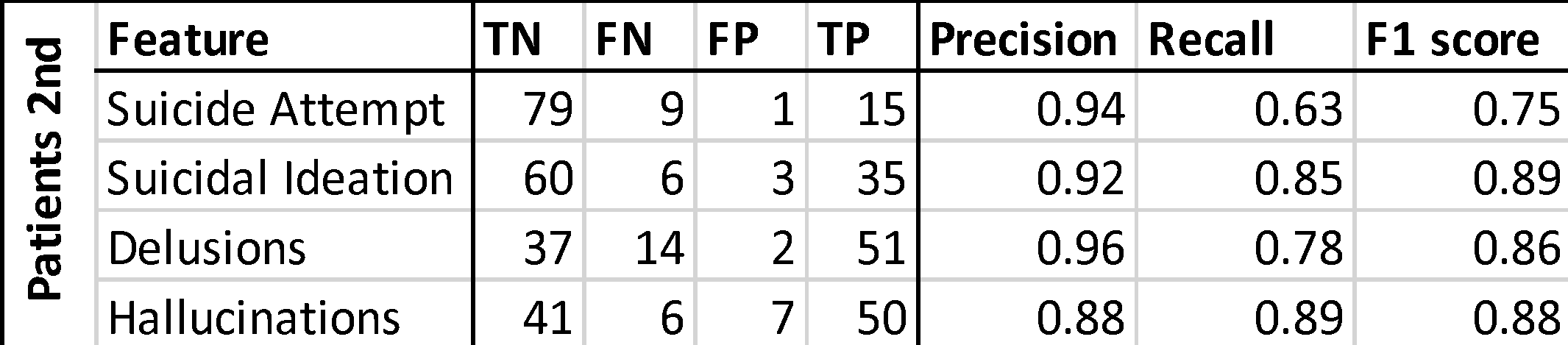

#### **Supplementary Table 8.** Demographic and clinical characteristics of the study cohort. Patients are classified by their most recent SMI diagnosis. Medians with interquartile range (IQR) are presented for: visits per patient, age at the most recent visit, length of stay, and length of the medical record. Tests comparing these values across the three main diagnoses (MDD, BD, and SCZ) are provided in the bottom part of the table. **Test*:** Across the three diagnoses, differences in percentages are tested with a chi-squared test, and differences in distributions are tested with a Kruskal-Wallis test. **Test**:** Between pairs of diagnoses, differences in percentages are tested with z-tests, and differences in distributions are tested with Mann-Whitney tests. Asterisks mark significant results at the Bonferroni-corrected alpha threshold of 0.05/8=0.006.

In this analysis, we observed distinct demographic and healthcare use patterns across diagnoses. Gender distribution showed that MDD and BD patients are predominantly female (over 60%), while SCZ patients are predominantly male (over 70%). Age of onset also varied, with MDD showing the earliest onset: almost 20% of MDD patients had their first visit before age 18, compared to only 12.6% of BD patients. However, at the time of their most recent visit, SCZ patients were the youngest group, averaging 33.7 years old, while BD patients were the oldest at 45 years old. Visits data show that BD and SCZ patients have more than double the number of visits in their records compared to MDD patients, with longer overall record durations. Hospitalization rates also differed significantly: over 70% of BD and SCZ patients were hospitalized at some point, compared to 56% of MDD patients. Lastly, SCZ patients had the longest average hospital stays at 14 days, while MDD patients had the shortest at 8 days.

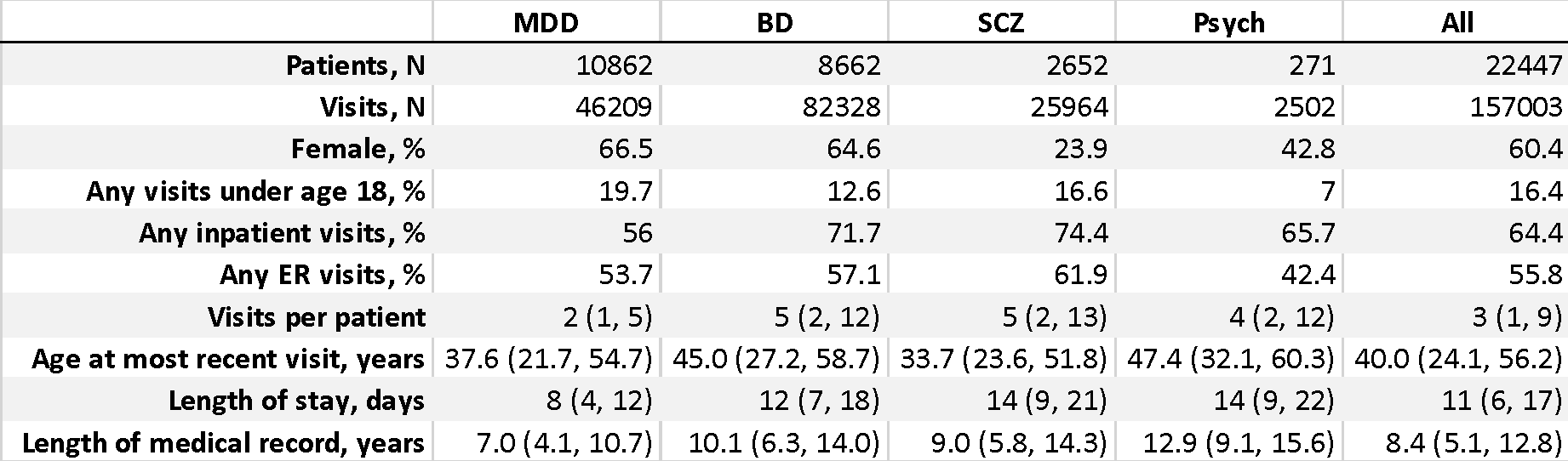

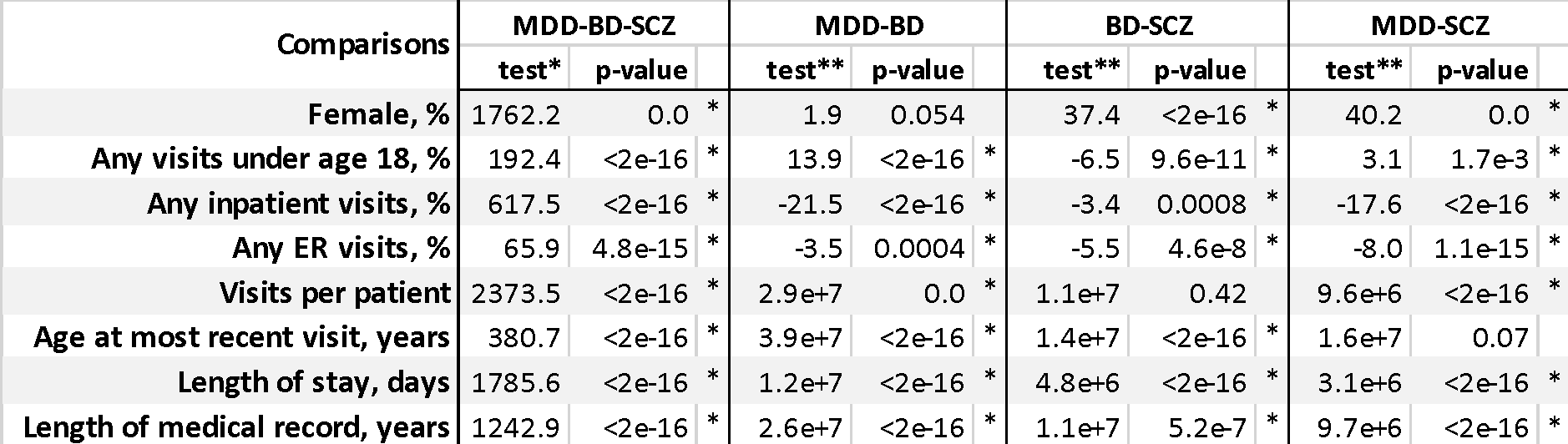

#### **Supplementary Table 9.** Frequency of each clinical feature (in percentages) for patients with SMI diagnoses, stratified by gender and inpatient history (yes: patients with a history of at least one inpatient hospitalization; no: individuals without any history of inpatient hospitalization). The first two columns show the total number of individuals included in this analysis, while the other columns show the frequencies of each clinical feature. The rows display the total numbers and frequencies considering all SMI diagnoses (“All”) and then considering each of the three diagnoses separately. All patients included in this table had at least two clinical notes in their EHR.

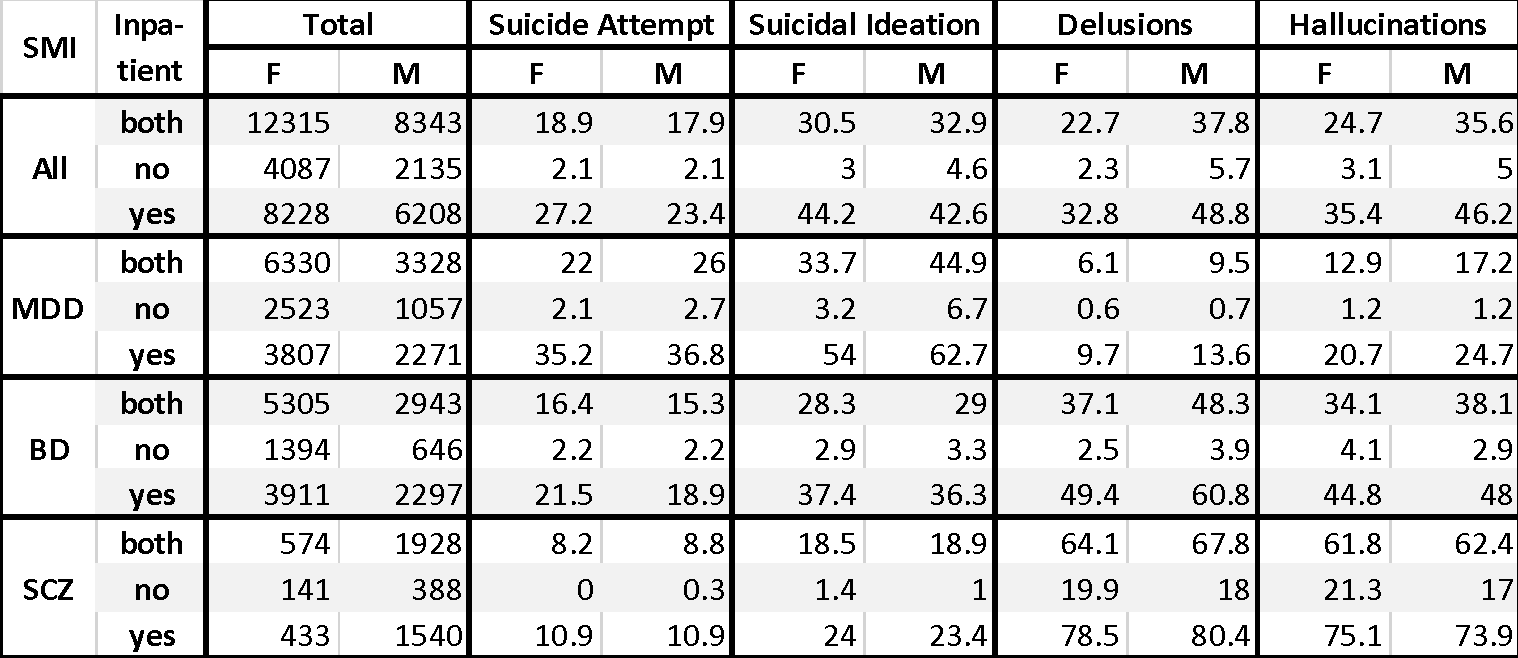

#### **Supplementary Table 10.** Odds ratios for patient-level associations of each clinical feature with gender, diagnosis, and other clinical features. Bonferroni-corrected alpha is 0.05/12=0.0041 for Table A and 0.05/16=0.0031 for Table B. Analyses are described in Supplementary Note 2.

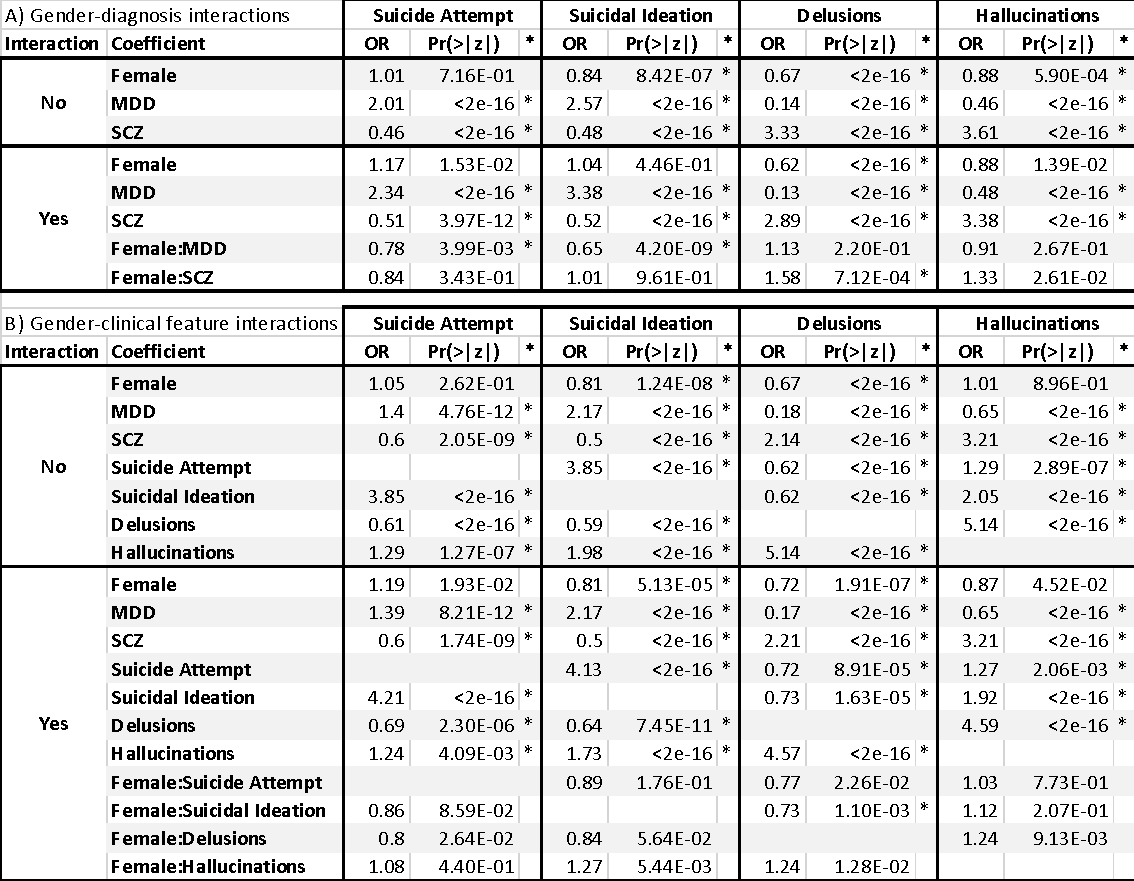

#### **Supplementary Table 11.** Percentage of individuals with comorbidities within each SMI diagnosis, as observed in patients with at least three encounters (n=12,962). The ICD codes for the 20 most frequent diagnoses are shown.

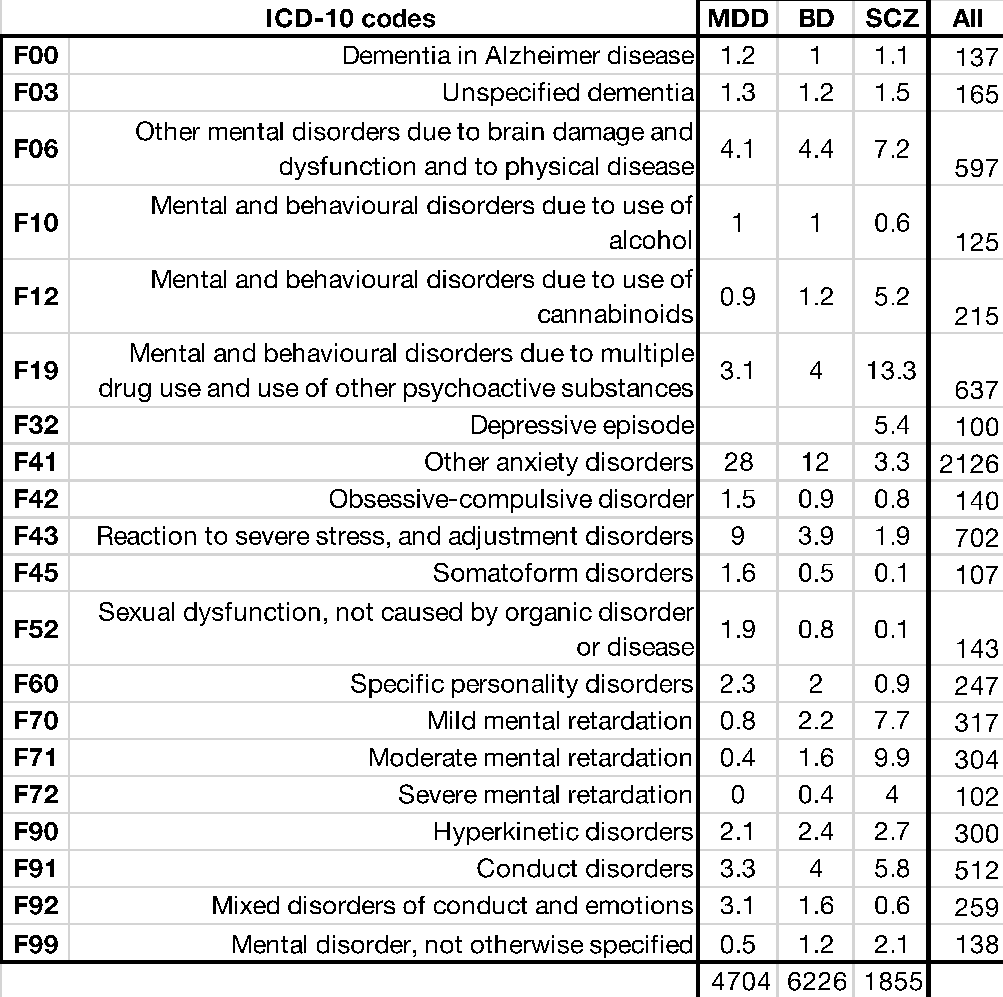

#### **Supplementary Table 12.** Most frequent diagnostic switches in individuals with high levels of diagnostic instability. These individuals are defined as having five or more diagnostic switches and at least one of them occurring after five years of illness.

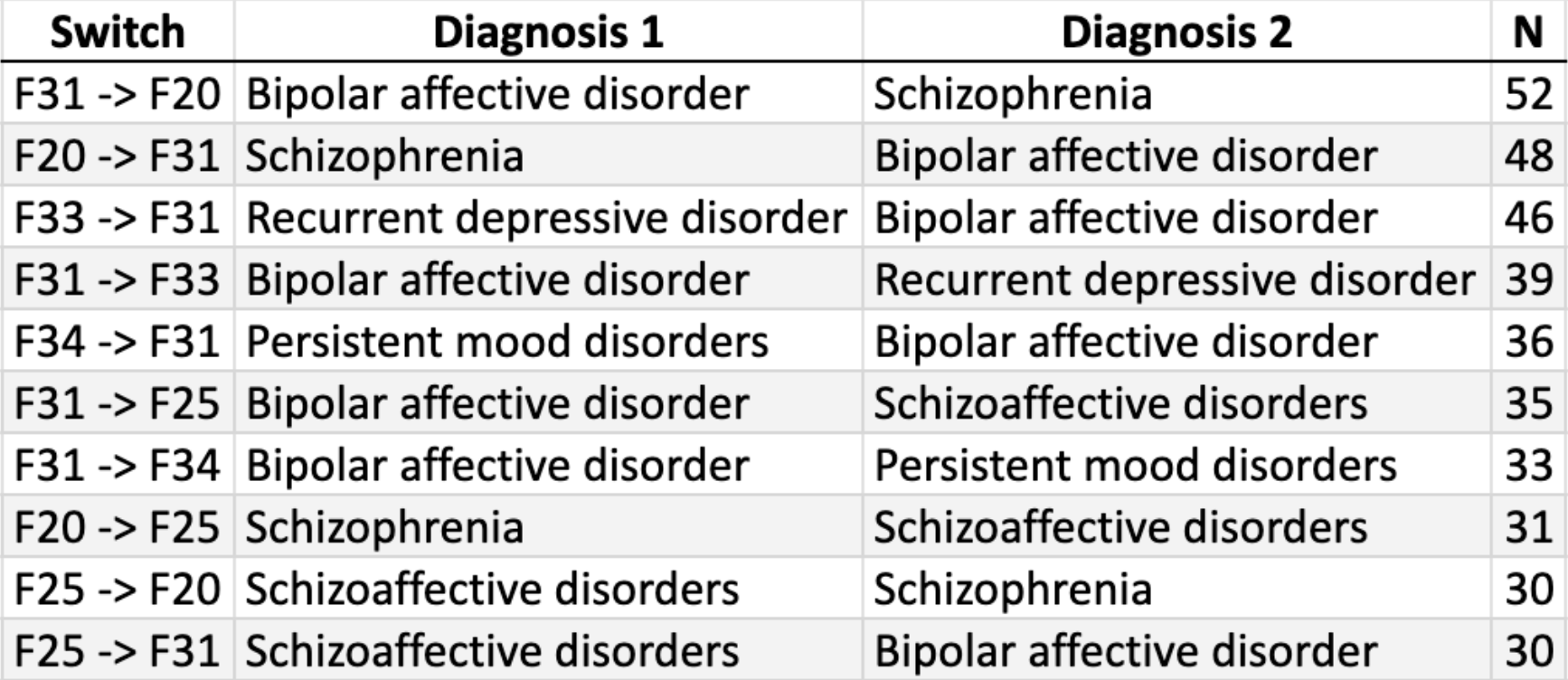
